# Task-voting for schizophrenia spectrum disorders prediction using machine learning across linguistic feature domains

**DOI:** 10.1101/2024.08.31.24312886

**Authors:** Rui He, Víctor Ortiz-García de la Foz, Luis Manuel Fernández Cacho, Philipp Homan, Iris Sommer, Rosa Ayesa-Arriola, Wolfram Hinzen

## Abstract

**Background and Hypothesis:** Identifying schizophrenia spectrum disorders (SSD) from spontaneous speech features is a key focus in computational psychiatry today.

**Study Design:** We present a task-voting procedure using different speech-elicitation tasks to predict SSD in Spanish, followed by ablation studies highlighting the roles of specific tasks and feature domains. Speech from five tasks was recorded from 92 subjects (49 with SSD and 41 controls). A total of 319 features were automatically extracted, from which 24 were pre-selected based on between-feature correlations and ANOVA F-values, covering acoustic-prosody, morphosyntax, and semantic similarity metrics.

**Study Results:** ExtraTrees-based classification using these features yielded an accuracy of 0.840 on hold-out data. Ablating picture descriptions impaired performance most, followed by story reading, retelling, and free speech. Removing morphosyntactic measures impaired performance most, followed by acoustic and semantic measures. Mixed-effect models suggested significant group differences on all 24 features. In SSD, speech patterns were slower and more variable temporally, while variations in pitch, amplitude, and sound intensity decreased. Semantic similarity between speech and prompts decreased, while minimal distances from embedding centroids to each word increased, and word-to-word similarity arrays became more predictable, all replicating patterns documented in other languages. Morphosyntactically, SSD patients used more first-person pronouns together with less third-person pronouns, and more punctuations and negations. Semantic metrics correlated with a range of positive symptoms, and multiple acoustic-prosodic features with negative symptoms.

**Conclusions:** This study highlights the importance of combining different speech tasks and features for SSD detection, and validates previously found patterns in psychosis for Spanish.

## Introduction

Human language is organized across a number of integrated levels, each of which carries a wealth of computationally extractable information relevant to mental health^1^, in a situation where diagnosis is still essentially based on information obtained from conversational speech. At the acoustic level, modulations in volume, pitch, speed of articulation, phonation-vs.-pausing times, and spectral properties of the speech sound can signal an underlying disease process, as can more monotonous speech at the level of prosody^2^. As language intrinsically carries meaning, the choice of words and their organization into meaningful sentences reflects the contents of thought generated in response to some tasks such as a picture description or answering a question^3^. Responding verbally to such tasks further adheres to formal-syntactic patterns or rules as reflected in the construction of phrases containing other phrases as projected from ‘parts of speech’ categories such as nouns, verbs, adpositions, determiners, and auxiliaries. Together, the complex edifice arising from integration across these levels of organization promises sensitivity to processes of mental change and decline.

Numerous studies employing machine learning with selected features across these speech and language domains have obtained classification accuracies for psychosis relative to healthy controls across several languages, ranging for the acoustic-prosodic domains from 76%^4^ to 94%^5^, despite the limitations of small sample sizes and lack of external validations. Features from spontaneous speech mostly performed well in classifying converters to psychosis from a high-risk state versus non-converters^6–8^ (but see Bianciardi et al.^9^), distinguishing different stages in the schizophrenia spectrum^10^, and predicting aspects of disease progression in longitudinal samples^11^. These studies have generally validated the usability of spontaneous speech as a digital biomarker for schizophrenia spectrum disorders (SSD).

These classification results are all obtained from data that are effortlessly produced and automatically analyzable given appropriate pipelines. Nonetheless, from a translational perspective, a number of desiderata currently stand out. First, studies have used a variety of speech elicitation tasks, from free conversational speech, picture or cartoon descriptions, dream reports, story recall, to reading and clinical interviews. As different tasks pose different cognitive demands, the effects of the task chosen are currently unclear and of potential clinical importance. Morgan et al. showed that free speech was less effective than picture description and story retelling in distinguishing groups of first episode of psychosis, clinical high risk, and controls^12^. Second, the relative importance of different feature domains for classification has only recently begun to be addressed in a small number of studies. Moshe et al. show through an ablation study that acoustic features capture more information than semantic ones extracted form text.^13^ Huang et al. found that acoustic features played a powerful role in classifying PANSS items featuring some relation to emotions, e.g., excitement, anxiety, emotional withdrawal^14^. Voppel et al. showed that combining a quantification of the semantic similarities of produced words with the word2vec language model and acoustic features extracted with openSmile boosted performance of a random forests classifier from 80 and 81%, respectively, to 85%^15^.

In this study, we developed a task-voting pipeline where identical machine learning classifiers were trained for SSD detection on five tasks with a comprehensive set of features from three linguistic domains to address the challenges with speech tasks and feature domains. To this we added further ablation studies to shed insight on the importance of different tasks and features.

## Methods

### Dataset

Ninety-two subjects were recruited in this study, including 41 healthy controls and 49 chronic SSD patients. As shown in Table 1, there was no significant difference between groups on age, sex, and years of education. Among them, eighty-eight are native Spanish speakers while one patient is native in French and another patient native in Portuguese. Both learned Spanish in school and speak Spanish fluently. Symptom severity of patients was evaluated with 10-item Positive and Negative Syndrome Scale (PANSS), including three positive items, three negative items, and four general items. Positive items include delusions (P1), conceptual disorganization (P2), and hallucinatory behavior (P3). Negative items included blunted affect (N1), passive/apathetic social withdrawal (N4), and lack of spontaneity and flow of conversation (N6). General items included anxiety (G2), mannerisms & posturing (G5), depression (G6), and unusual thought content (G9).

**Table 1.** Demographics and clinical scores of the dataset.

|  | Control | Schizophrenia | Test | Statistics | *p* value | Effect size |
| --- | --- | --- | --- | --- | --- | --- |
| Number | 41 | 49 | / | / | / | / |
| Age | 41.79 ± 7.79 | 39.93 ± 9.61 | *T* test | 1.012 | 0.314 | 0.210 |
| Sex | 41.46% | 48.98% | X<sup>2</sup> test | 0.251 | 0.617 | 0.053 |
| Education | 11.56 ± 2.57 | 10.94 ± 3.50 | *T* test | 0.967 | 0.336 | 0.201 |
| PANSSP | / | 3.0 (2.0) | / | / | / | / |
| PANSSN | / | 4.0 (2.2) | / | / | / | / |
| PANSSG | / | 5.0 (3.0) | / | / | / | / |
| PANSSTotal | / | 13.0 (4.2) | / | / | / | / |

Note: Age and education are indicated by mean ± standard deviation by years. Sex is indicated by the proportion of female participants. PANSS scores are indicated by the median and interquartile range (IQR). We used the 10-item PANSS version in this study (P1, P2, P3, N1, N4, N6, G2, G5, G6, and G9). PANSSP indicates the median (IQR) of the total sum scores of 3 positive items. PANSSN indicates the median (IQR) of the total sum scores of 3 negative items. PANSSG indicates the median (IQR) of the total sum scores of 4 general items. PANSSTotal indicates the median (IQR) of the total sum scores of all 10 items.

### Speech elicitation and transcription

Every participant went through the DISCOURSE in Psychosis Speech Collection Protocol as available at https://discourseinpsychosis.org/resources/. Audio was recorded using Audacity with manual tagging to isolate participant speech at Valdecilla Biomedical Research Institute (IDIVAL). The protocol and methodology for this study was approved by the local research ethics committee (CEIm internal code 2021.119). We extracted five audios from the three tasks in total: (1) Self-related interview (SelfInt): free speech elicitation on topics related to the interviewees, e.g. *Tell me a bit about yourself (¿Puedes hablarme un poco de ti?)*; (2) Past-related interview (PastInt): free speech on topics related to events that happened in the past or in recent years, e.g. *Thinking back, can you tell me a story about something important that happened to you in your life? (Haciendo memoria, ¿puede contarme una historia sobre algo importante que le haya ocurrido en su vida?)*; (3) Picture description (PicDesp): describe three pictures from Thematic Apperception Test (TAT); and (4) story recall: read a story and retell it immediately without looking at it. Recordings of reading and retelling were separately analyzed. All speech was transcribed with Whisper^16^ and manually checked and corrected by a researcher. For both SelfInt and PastInt, we concatenated the answers into one paragraph. For PicDesp and retelling, we treated each description of a picture and the retelling as single paragraphs. We did not include the transcripts of the reading part.

### Acoustic-prosodic analysis

For each task, we concatenated the interviewer’s voice into one recording and thus obtained five recordings for each participant. We first used OpenSmile to extract 88 features from the eGeMAPSv02 feature set, which was developed as a minimalistic parameter set for acoustic computing including voice quality, pitch profile, and spectral and cepstral coefficients^17^. In addition, to have a more detailed and comprehensive understanding of the prosodic profile, we used Prosogram to extract 31 additional prosodic features describing the quantity of speech and pitch variations^18^. Details on the prosodic variables can be found in Supplementary Table S1.

### Morphosyntactic complexity

Speech transcripts were split into sentences and words with the Spanish model from SpaCy. We first extracted six quantity features, including the number of words, the number of words excluding stopwords, the number of sentences, the number of noun chunks, the type token ratio of words, and the ratio of stopwords. The spaCy model also tagged word classes and analyzed relevant morphological changes. With these results, we computed the ratio of the 16 universal part-of-speech tags and 18 morphological features, including the ratio of four tenses, the ratio of three different personal pronouns, the ratio of negation, the ratio of four moods, the ratio of masculine and feminine nouns, the ratio of singular and plural nouns, and the ratio of definite and indefinite nouns. In addition, we parsed the sentences into dependency and constituency structures to extract formal syntactic features, which has been shown to change in SSD.^3,19^ Dependency parsing was performed using spaCy’s dependency parser to identify pairs of words with direct dependency relationships. For example, in the sentence *She ate an apple, an* is the determiner (det) of *apple*, and *She* is the nominal subject (nsubj) of *is*. The ratios of 26 dependency relationships were extracted. Distance between two dependent words, as measured by the number of intervening words, is a well-established indicator of syntactic complexity and cognitive effort.^20,21^ Both the average and maximum dependency distances were thus calculated to index dependency complexity. Another way to parse *She ate an apple* is by examining its hierarchical phrasal structure: *an apple* is a noun phrase embedded in the verb phrase *ate an apple*, which, along with the noun phrase *She*, forms the complete sentence, as in *[[She]*_*NP*_ *[ate [an apple]*_*NP*_]_*VP*_]_*S*_. The phrasal structures were identified with a Spanish constituency parser from Stanza, and represented as directed acyclic graphs. From the graphs, we computed the distance between each node and the sentence node as the count of intervening nodes to identify the syntactic depth of each word node, and extracted eight relevant features to index the constituency complexity. In total we extracted 76 morphosyntactic features and the details can be found in Supplementary Table S2.

### Semantic similarity analysis

Given a text split into *N* units {*U*_1_,*U*_2_,…, *U*_*N*_}, we vectorized each unit with certain language model into a matrix of embeddings {*e*_1_,*e*_2_,…, *e*_*N*_}. The similarity between vectors *e*_*i*_ and *e*_*j*_ was defined by the cosine value of the angle between them:

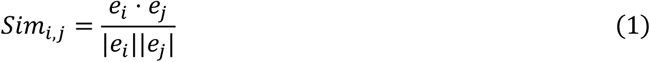

where |*e*_*i*_ | and |*e*_*j*_ | are the norms of the vectors e_i_ and e_j_, respectively.

We first computed the averaged cosine similarity between adjacent words as first-order mean similarity (MeanK1):

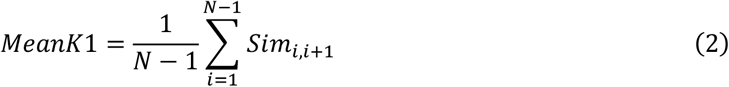

Then, the second-order mean similarity was defined as the averaged cosine similarity between two units with one unit in between (MeanK2):

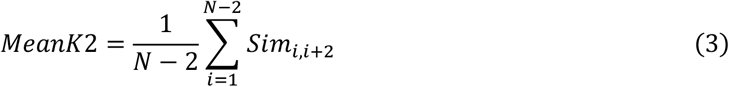

Global semantic similarity was defined as the averaged cosine similarity between all unit pairs:

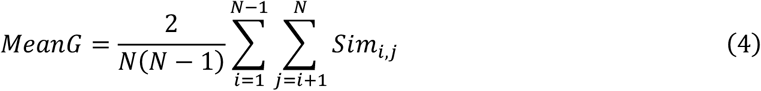

In addition to these averaged scores, which previous studies have often found indicative of semantic changes in speech in SSD,^3,22,23^ we introduced two sets of variables to approximate the dynamics of a navigation in semantic space. Given a time series of semantic similarity between adjacent units {*Sim*_1,2_,*Sim*_2,3_,…, *Sim*_*N*−1,*N*_}, one set comprised six statistical measures describing the distribution of the similarities, including variance, maximum (peak), minimum (valley), amplitude (i.e. difference between maximum and minimum), skewness, and excess kurtosis (i.e. kurtosis – 3, where 3 is the kurtosis of normal distribution). Another set of variables took the temporal order of the sequence into account and aimed to depict the temporal dynamics with six variables. Let x_i_ represent the *i*th semantic similarity in the array {*S im*_1,2_,*Sim*_2,3_,…, *Sim*_*N*−1,*N*_} and 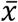 represent the mean value of this array (i.e. MeanK1). Then the mean crossing rate (MCR) measures how frequently a semantic similarity score crosses its mean value 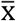:

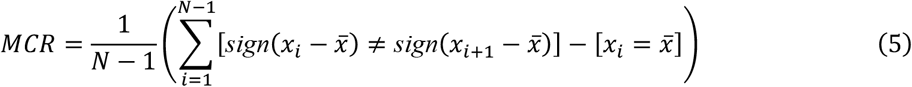

Slope sign changes (SSC) measures the normalized number of times the slope of the signal changes its sign:

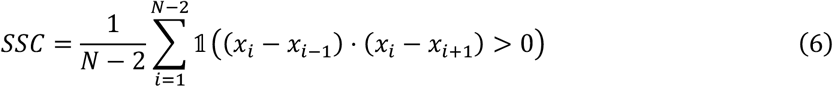

where 𝟙 is an indicator function that returns 1 if the condition inside is true, and 0 otherwise.

The wave length (WL) calculates the average absolute difference between two consecutive semantic similarity scores:

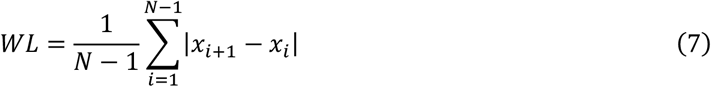

Approximate entropy (ApEn) measures the unpredictability of fluctuations over time. Higher ApEn indicates a more predicable time series with increasing amount of regularity in its fluctuations. ApEn was estimated using the Python package called *Antropy* with default parameters.

Autocorrelation function (ACF) computes the correlation coefficients between the time series and copies of itself that are temporally shifted with a series of lags:

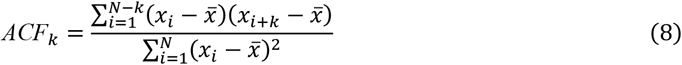

where *k* is the number of lags. We extracted ACF with one lag (ACF_1_) and zero crossing rate of the ACF waveform (AcfZcr).

Only text with over four units entered the semantic analysis. Text less than four units was treated as missing values.

### Semantic embedding retrieval

We utilized three different language models to proxy semantic structure at different levels. After removing all punctuations and spaces (as defined by the part-of-speech tags from SpaCy) and stopwords (as defined in the nltk package), all tokens were encoded with the fastText model pretrained on Spanish data.^24^ FastText is a context-free model assigning the same embedding to the same token regardless of the actual context, while contextual models like transformer-based models can encode the tokens with the context taken into account. We applied roberta-large-bne, a transformer-based language model pretrained on Spanish corpora,^25^ to tokenize the sentences and encode every token into embeddings. Finally, at the sentence level, we applied a Spanish sentence-transformers model to compare the similarity between two sentences (hiiamsid/sentence_similarity_spanish_es).

### Semantic centroid analysis

Following Xu et al., we computed the similarity not only among adjacent units, but also between the units and their centroid.^26^ Given a listing a matrix of embeddings {*e*_1_,*e*_2_,…, *e*_*N*_}, the static centroid was defined as the averaged embedding across all units:

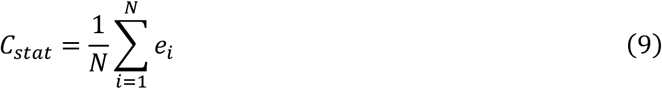

The cumulative centroid was defined as the averaged embedding over all preceding units. The cumulative centroid at the *j*th unit is thus:

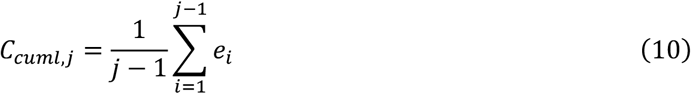

The centroid serves as a proxy of the central ‘topic’ of the text as a whole. While the static centroid summarizes the whole text, the cumulative centroid may reveal how the topic evolves as the narrative unfolds. We computed the similarity between every unit and the centroid, whether static or cumulative, as a time series from the first unit to the last unit. Then, we extracted the MeanK1, six distribution variables and six dynamic variables as described above.

### Prompt to response similarity

As all speech data were elicited by prompts, either verbal or visual, analyzing the similarity between prompts and responses can indicate how much speakers deviate from the trigger or external ‘anchor’ of their speech. For this purpose, for the interview tasks, the texts were no longer concatenated, but divided into several ‘chunks’, where each chunk contains several words from the interviewer (the prompt) and another piece of text from the interviewee (the response). We used the Spanish sentence-transformers model to compare sentence similarity between each prompt and response per chunk and then averaged across chunks for each participant. For story reading and retell, we used the Spanish sentence-transformers for the similarity between the original and recalled story. For picture descriptions, we used the multimodal CLIP model, which can encode the visual prompt and elicited speech at the same time to enable similarity comparisons between them two^27^.

### Identify SSD with different tasks as ‘voters’

In total, 319 linguistic features were extracted for each participant from each of the five recordings and their corresponding transcripts: SelfInt, PastInt, PicDesp, Read, and Retell. For PicDesp, semantic and morphosyntactic features were averaged across three pictures. All features were first z-scored within each task, and missing values were filled with the minimum values per feature. For story reading, semantic and morphosyntactic features were all filled with zeros. We trained five Extra Trees classifiers on the five tasks for SSD identification (parameters: n_estimators: 5, max_depth: 5, min_samples_split: 5, min_samples_leaf: 2, max_features: sqrt, criterion: entropy, random_state: 42). Each classifier was trained to detect SSD from controls based on the linguistic features extracted from the five tasks, and they voted together for the final prediction. The final prediction was determined by majority voting: for example, if a participant received four predictions as SSD and one as control, they would be classified as SSD. We performed a nine-fold cross-validation on 90% of the data, with a hold-out dataset of 10% to evaluate model performance and generalizability. The cross-validation set itself was split into nine folds. Iteratively, in each cross-validation, the model was trained on eight folds, tested on the remaining fold, and also evaluated on the hold-out dataset. This process yields nine performance matrices per fold, which were then averaged into the cross-validation and hold-out performance matrices. Thus, the model never saw the hold-out dataset during training and validation. Performance was evaluated using precision, recall, accuracy, and F1-score.

### Feature selection

A large and comprehensive feature set as used here, though allowing for comprehensive characterization of the speech profiles, may contribute noise to the classifier and increase difficulty in interpretation. Selecting the most informative features to be forwarded to the classifier is therefore necessary and beneficial. We first removed features highly correlated with each other. Removing these features helps mitigate redundancy and multicollinearity, thereby improving interpretability and generalizability. We conducted repeated measures correlations (rmcorr) for each feature against others. Features showing strong correlations, as indicated by coefficients exceeding 0.9, were subsequently removed with only the first one remaining. Features were removed on first found, first removed basis. Forty-five features were excluded at this stage.

Following the removal of strongly correlated features, mixed-effect ANOVA was employed to gauge the discriminative power of each linguistic feature in identifying SDD. Features were ranked based on their ANOVA F-values, sorted from highest to lowest. We selected the *N* features with highest ANOVA F-values. The selection of the optimal number *N* was guided by an aggregated accuracy score criterion, defined as the sum of cross-validation and hold-out accuracy scores when their difference was less than 0.05, or divided by the difference when greater.

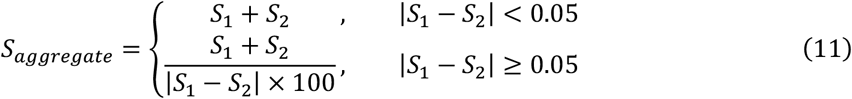

where *S*_1_ is the cross-validation accuracy score and *S*_2_ is the hold-out accuracy score. The model reached the highest aggregated accuracy score with 24 features.

### Ablation studies

To understand the importance of different linguistic domains and different tasks, we investigated the performance of the identical system after removing each task, and reported the difference as compared to the performance with five tasks on the hold-out set. Similarly, we investigated the performance of the identical system when removing each feature, averaged the difference for each of the three feature domains, and reported the averaged difference as compared to the performance with all selected features.

### Exploratory factor analysis

An exploratory factor analysis (EFA) was conducted to further understand the underlying structure of the selected features. A parallel analysis based on principle component analysis (PCA) suggested to retain five factors.^28^ We fit the data with a factor analysis model using minimal residual and promax oblique rotation. Details can be found in the supplementary materials.

### Group comparisons and symptom severity correlations

Finally, for each of the 24 selected features, we constructed a mixed-effect linear regression model to investigate the effect of diagnostic group (i.e. SSD or controls) on the feature. The model included fixed effects of age, gender, education years, and diagnostic group, and random effects of participant and task. All *p* values were corrected with False Discovery Rate (FDR) and reported as *q* values. We carried out additional mixed-effect models on other variables of interest as supplements. For example, we analyzed changes in proportion of pauses as a supplement to findings about changes in the duration of pauses. In addition, within the SSD group, we averaged the features across all tasks, for each participant, and performed Spearman’s correlations between the averaged features and PANSS items. FDR corrections were performed for each feature across the ten PANSS items.

## Results

### Model performance and ablation studies

With the 24 selected features, respectively on the cross-validation and hold-out sets, the model achieved precision scores 0.824 and 0.891, recall scores of 0.861 and 0.822, F1 scores of 0.832 and 0.846, and accuracy scores of 0.815 and 0.840. These 24 features included 16 acoustic features, 4 semantic features, and 4 morphosyntactic features, as reported in Table 2 with their ANOVA F-values. As shown in Figure 1A, removing each of the tasks led to slight increases on recall scores but greater decreases on other scores. Removing picture descriptionSimpaired the performance most, followed by story reading and then retelling, and then free speech about self and then about past events. Removing any of the features reduced the model performance, with the greatest decline seen when removing mean syntactic depth (accuracy decrease by 0.198), and the slightest decline seen when removing median nucleus duration (accuracy decrease by 0.049). As for feature domains, as shown in Figure 1B, removing morphosyntactic measureSimpaired the performance most, followed by acoustic measures and then semantic measures.

**Table 2.**
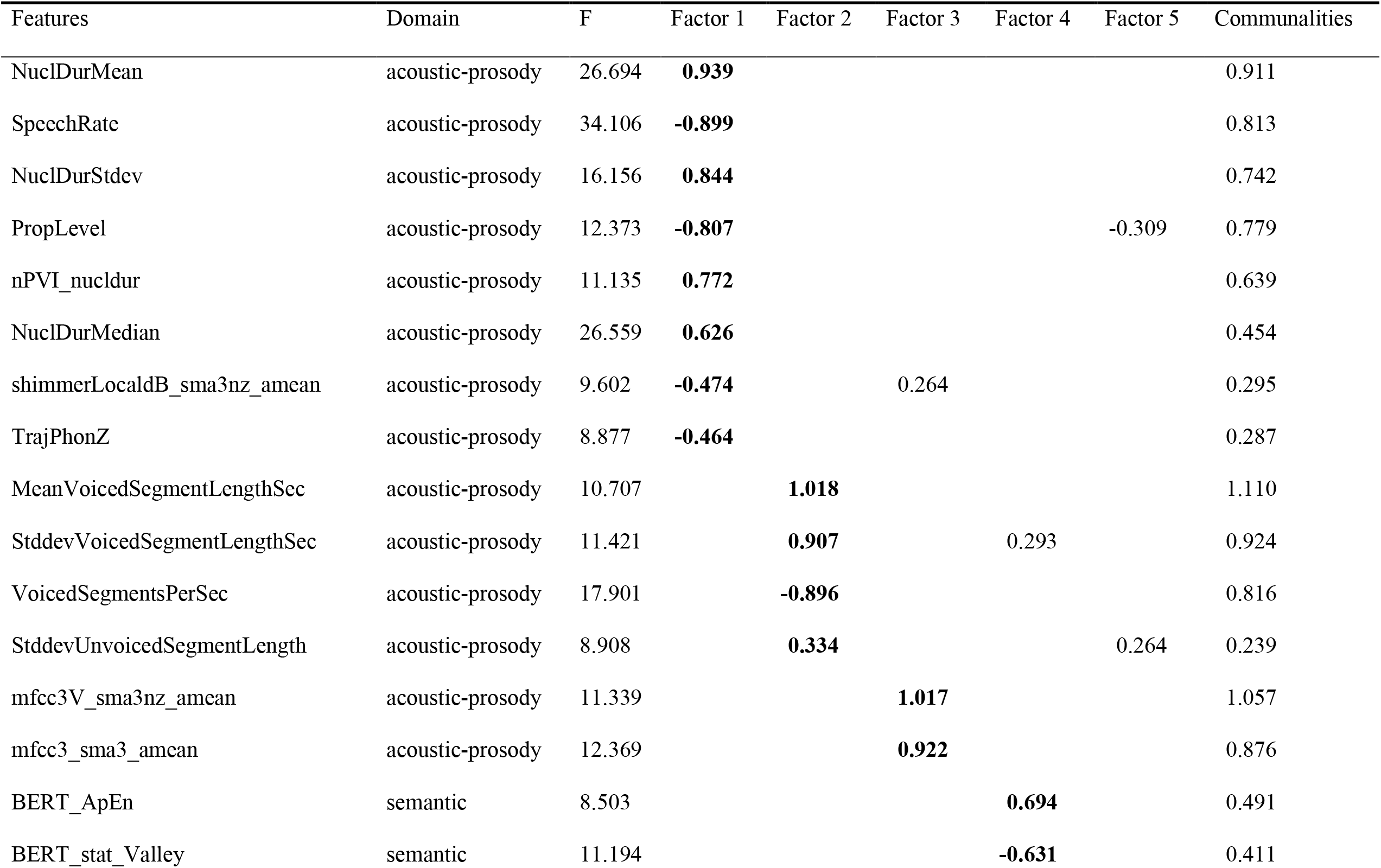
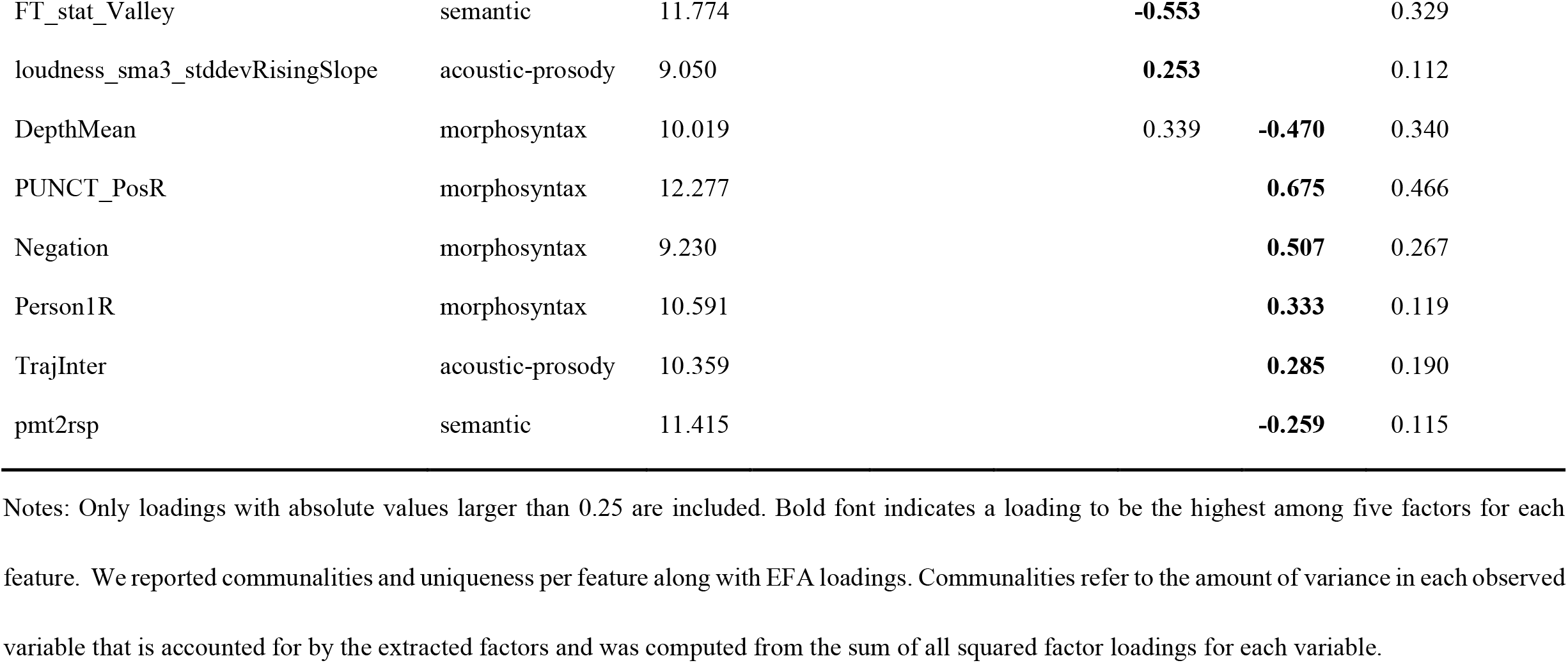
ANOVA F values and EFA factor loadings.

**Figure 1.**
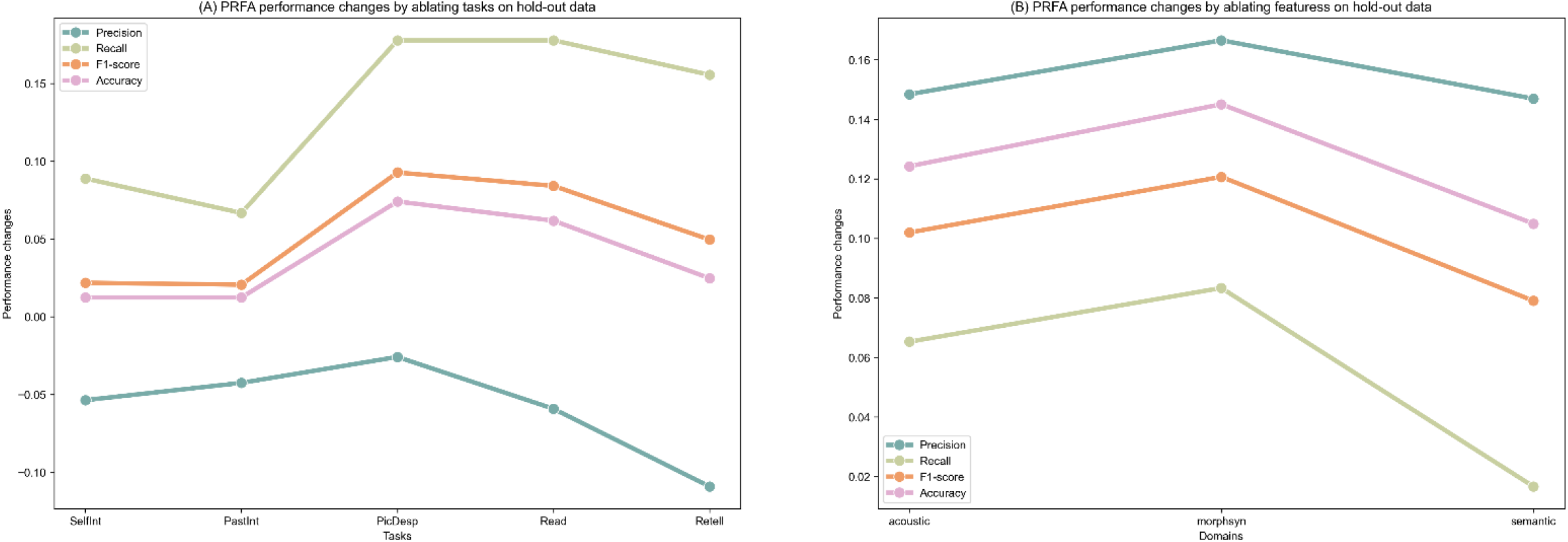
Ablation results on different speech tasks and feature domains. The changes in performance are defined by subtracting the performance of the ablated model from the performance of the whole model and showed on the y axis. (A) Changes in model performance by ablating each task. (B) Changes in model performance by ablating features from different domains.

### Factor analysis

Five factors were observed from the selected 24 features, as shown in Table 2. The first factor mainly includes speech quantity measures, together with one voice quality measure regarding shimmer and one pitch variation measure. The second factor concerns pauses in speech, while the third factor concerns MFCC values, and the fourth factor includes three measures derived from semantic similarity waves and one measure on loudness. Finally, the fifth factor includes all morphosyntactic measures, similarity between the speech and speech prompts, and one pitch variation measure.

### Group differences in selected features

The mixed-effect models suggested significant group differences on all selected features (*q* < 0.05). Results are visualized in Figure 2 and details such as *Z* and *q* values can be found in the Supplementary Table 4. In addition to these 24 measures, we also reported some additional measures to contextualize the speech changes with *Z* and *p* values reported here.

**Figure 2.**
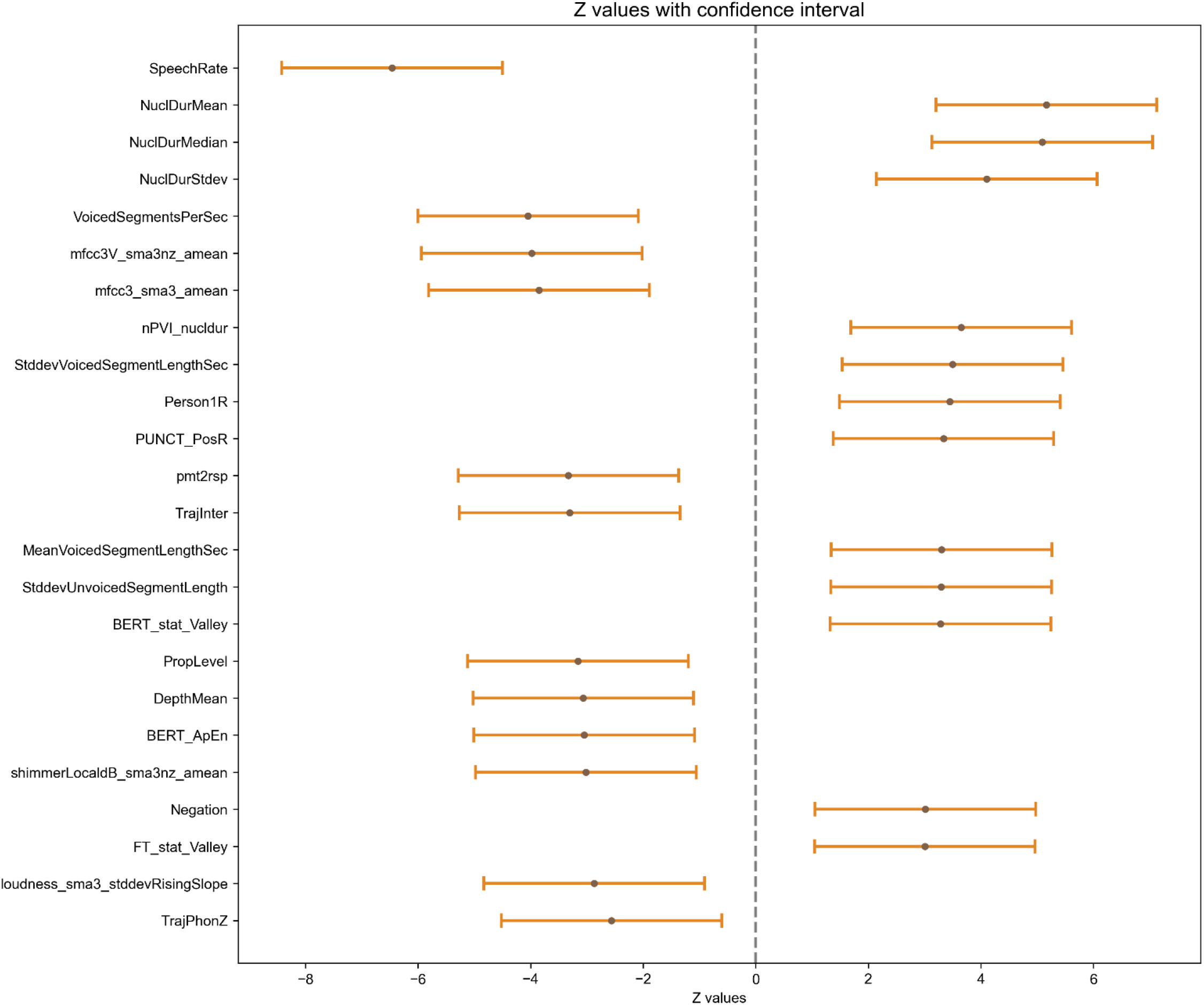
Results of mixed-effect regression model on the preselected variables. The error bars show the 95% confidence interval of the Z value (the central dot) for every feature forwarded to the classifier after feature selection. Features are listed on the y axis. The dash line indicates the position of 0. The error bars on the right side of the dash line indicate increases in SSD while those on the left indicate decreases in SSD.

The SSD group spoke more slowly, thus producing longer voiced speech, longer nucleus durations and longer pauses. However, the increase in the proportion of pauses was not significant (*Z* = 1.863, *p* = 0.063). In addition to an increase in general length, the variabilities in the durations of both voiced and unvoiced (i.e. paused) segments also increased. On the other hand, the variations in pitch, amplitude, and sound intensity decreased in SSD, as evidenced by decreases in pitch trajectories, higher proportion of nuclei without glissando, less shimmer (changes in amplitude), and lower standard deviation of the slope of rising signal parts of loudness. Lower MFCC coefficients indicate less energy or intensity in certain frequency ranges of the speech signal.

The most prominent change in the semantic domain was a decrease in the similarity between the speech and its prompt in SSD compared to controls. In addition, the minimal distance between the embedding centroid to each word increased in SSD, as derived from both fastText and BERT, but insignificantly decreased with sentences (*Z* = -0.744, *p* = 0.457). As for word-to-word similarity arrays, these became more predictable with BERT, also significantly with fastText (*Z* = -2.128, *p* = 0.033) and insignificantly with sentences (*Z* = -0.337, *p* = 0.736).

In the morphosyntactic domain, more first-person pronouns were used in SSD, together with insignificantly less second-person pronouns (*Z* = -0.427, *p* = 0.669) and significantly less third-person pronouns (*Z* = - 2.839, *p* = 0.005). We also observed more punctuations and more negations in SSD. The mean syntactic depth averaged across words decreased in SSD, but as a probable consequence of less words produced (*Z* = -2.006, *p* = 0.045). When normalized by sentence length, syntactic depth insignificantly *increased* in SSD (*Z* = 0.284, *p* = 0.777).

### Correlations with clinical symptoms

Spearman’s correlations between selected linguistic features and ten PANSS items were visualized in Figure 3 and details can be found in the Supplementary Table 4. No significant correlations were found with general items. Two semantic measures correlated with positive symptoms: Higher score in delusions correlated with lower similarity between prompt and response and higher minimal similarity between word embeddings and the static centroid from BERT. The former (lower similarity between prompt and response) also correlated with higher score in conceptual disorganization. The latter (higher minimal similarity between word embeddings and the static centroid from BERT) also correlated with hallucinatory behavior, as well as two negative symptoms, blunted affect and lack of spontaneity and flow of conversation. These two negative symptoms further correlated with multiple acoustic-prosodic features, including slower speech with longer nucleus durations and longer pauses, and less variations in pitch trajectory and sound intensity.

**Figure 3.**
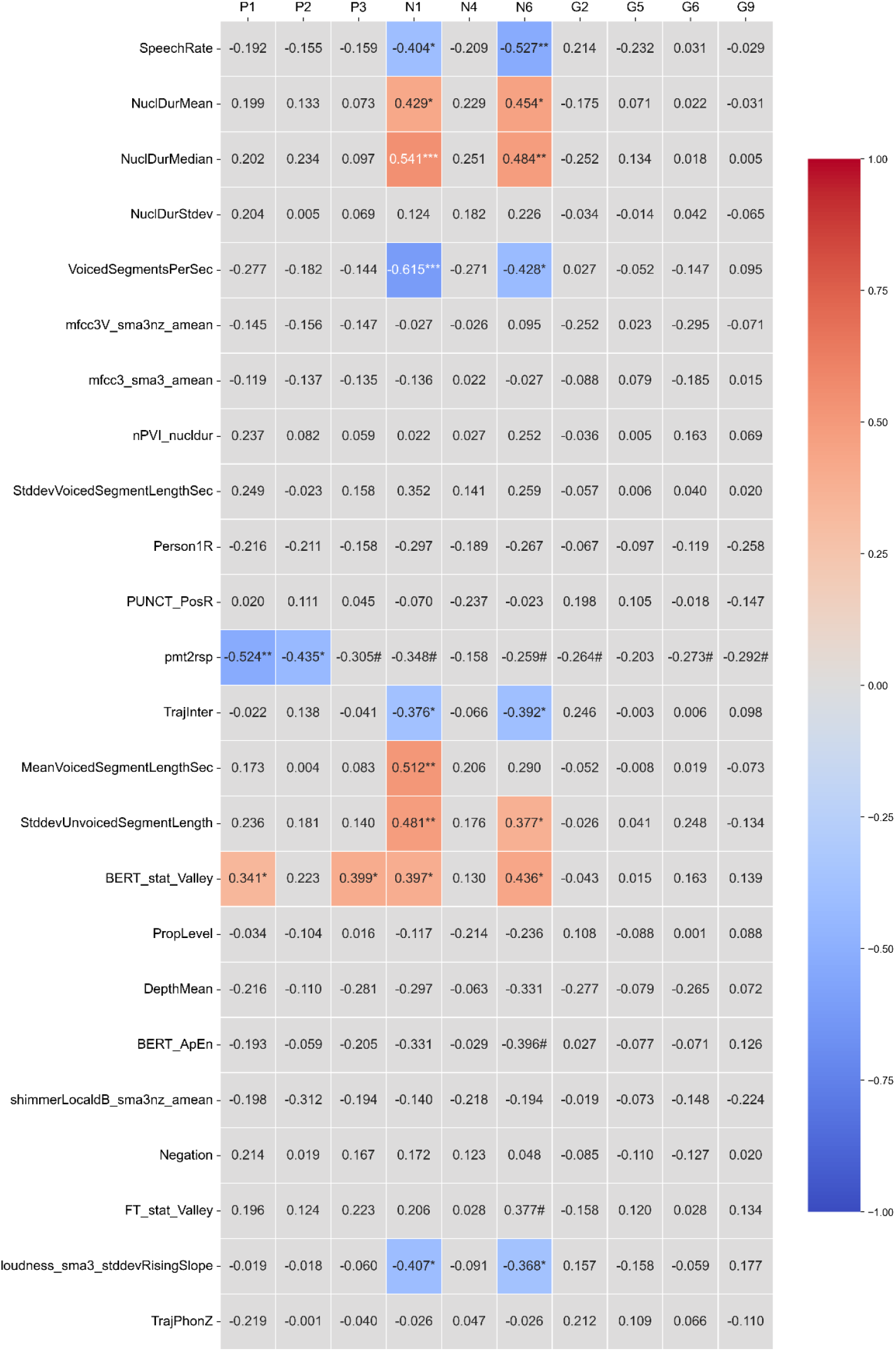
Correlations between selected features (on the y axis) with the PANSS items (on the x axis). Only significant correlations were colored. The numbers indicated the correlation coefficients. Positive correlations were highlighted with warm color while the negative correlations were highlighted with cold color. The density of the color increased with the strength of correlation. * *q* < 0.05, ** *q* < 0.01.

## Discussion

This study proposed a new model of combing information from different language tasks to let them vote for SSD classification. We showed that combining various speech elicitation tasks with features from multiple linguistic domains enhances the classification performance of SSD versus healthy controls. While omitting any task might slightly improve precision, it results in greater reductions in recall, as well as in accuracy and F1 scores. Removing any feature adversely affects the model’s performance across all four metrics. The satisfactory performance scores (precision: 0.891, recall: 0.822, F1 score: 0.846, accuracy: 0.840 on the hold-out set) confirm the diagnostic and prognostic potential of spontaneous speech, proposed as a digital marker^29^, or even a mechanistic factor^30^, for psychosis. This level of performance was achieved not only on the cross-validation set but also on unseen hold-out data, indicating the model’s generalizability.

Mixed-effect regression models on the selected variables indicated that the model learned the following speech patterns to detect SSD: SSD patients spoke slower with higher variations in temporal features, while variations in pitch, amplitude, and sound intensity decreased. Minimal semantic distances between embedding centroids and each word increased, while word-to-word similarity arrays became more predictable in SSD^3^. Again consistently with previous studies^3,39^ this pattern of a contracted semantic space does not exclude and is indeed consistent with a decrease in the semantic similarity between speech and prompts when using bimodal models, showing greater deviance (higher semantic distance) of speech from its external ‘anchor’ in SSD.

In SSD, a wide range of speech tasks has been employed to observe abnormal patterns in speech. While describing TAT pictures has been widely adopted^3,31,32^, other spontaneous speech tasks, such as free speech^33^ and story recall^34^, have also been used for speech elicitation. Comparisons of different tasks in the capacity of distinguishing patients from neurotypical populations have been investigated in other diseases such as the Alzheimer’s disease (AD) spectrum. Recall tasks have been reported to be most efficient in detecting AD as compared to picture description and conversation, potentially due to the memory component involved in this task^35–37^. Task difference, however, is underexploited in SSD, though Morgan et al. found that recall and picture description outperformed free speech^12^. While all five tasks contributed to the model performance, the TAT-based picture description task made the greatest contribution, underscoring the usability of this commonly employed task. Story reading and recall, which has scored high in AD detection, also made substantial contributions, though less than the picture description tasks. Meanwhile free speech contributed the least to the voting classifier, but removing it still worsened the model’s performance. Overall, these results suggest that, in a real-world clinical setting, clinicians should employ a comprehensive set of speech tasks to maximize the information available for SSD classification. Morphosyntactically, more first-person pronouns were used together with less third-person pronouns, and more punctuations and negations.

Compared to task choice, it is more often noted that different feature domains contribute differently to SSD detection^13,15,38^. Our results confirm initial findings of Voppel et al, showing that a holistic linguistic profile, with acoustic-prosodic, morphosyntactic, and semantic structure features classifying together, empowers the model for best performance. Every feature mattered to the classification, as removing any one of them would lower the performance. However, unlike previous findings showing that acoustic-prosodic features were more powerful in classification than textual measures^13,15,38^, the ablation results revealed that removing morphosyntactic measures, particularly mean syntactic depth, was most detrimental, followed by acoustic-prosodic and semantic measures. Our exploratory factor analysis offers a potential explanation for this inconsistency. It identified five latent factors underlying the 24 selected features. Speech quantity, pauses, MFCC, and syntactic features were distinctly separated into different factors. In contrast, semantic measures, as well as a few voice quality and pitch measures, were distributed *across* different factors, suggesting a greater degree of interrelations with other measures. Specifically, semantic similarity measures were intertwined with loudness changes and mean syntactic depth, while prompt-response similarity was connected with syntactic measures and pitch trajectory. This suggests that when removing a highly interrelated measure, the model can still capture some aspects of it through related features, such as syntactic measures for prompt-response similarity, though not as effectively as it would with the feature itself. Conceivably, ablating semantic measures for this reason made the least impact on model performance, whereas removing morphosyntactic measures caused the highest impairment. From this point of view, our findings do not necessarily contradict previous research but rather provide complementary evidence regarding the importance of different feature domains.

The mixed-effect regression models carried out on all selected features, which included fixed effects of age, gender, education, and diagnostic group, contribute to model interpretation. While we replicated a previously documented acoustic and semantic pattern as discussed above, another morphosyntactic pattern rather extends a varied picture seen previously: more first-person pronouns were used together with less third-person pronouns, and more punctuations and negations. The first of these results contradicts a previous study^40^ finding fewer first-person singular pronouns in SSD, while another study found more^41^. Use of first person is likely to be highly task-sensitive, with picture descriptions in particular not inviting first person use, and with free speech questions doing so to various degrees. Crosslinguistic differences may also matter to variability between studies, including whether a language marks grammatical person primarily on pronouns or in verbal morphology, and whether NLP tools used to detect first or third person or sensitive to both of these. Apart from these factors, it is also possible that use of first person is simply too individually variable in ordinary language use to form a promising part of a crosslinguistically replicable pattern in SSD.

As for the relations to symptom severity, acoustic-prosodic features were found to correlate only with negative symptoms including blunted affect (N1) and lack of spontaneity and flow of conversation (N6). Higher severity in these two negative symptoms were indicated by slower and prolonged speech with more pauses and less pitch variations. This apriori predictable pattern lends further confidence to tracking negative symptomatology through these acoustic and prosodic features. Contraction in the semantic space and deviance from the speech prompt were also indicative for these two negative symptoms. Moreover, these two semantic measures were the only measures correlated to positive symptoms. Specifically, minimal distances between words and the text centroids (averaged embeddings from BERT of the texts they belong to) correlated with delusions and conceptual disorganization, while prompt-to-response similarity correlated with delusions and hallucinations. Speech changes, especially poverty of speech, has often been thought of as one of the clearest biomarkers for negative symptoms^42–45^. Our results raise the expectation that, the semantic structure of speech may particularly capture changes in positive symptoms, which may be highly valuable in the context of tracking a patient’s transition into remission, where positive symptoms recede, and a transition back to psychosis (relapse), where they re-emerge.

Together, our study proposes a new classification model for speech-based SSD detection. Combining different speech tasks and linguistic domains can effectively elevate model performance. Our results should be considered as being subject to several limitations. First, our study had a relatively small sample size. Machine learning models on small datasets usually face high risk of bad performance and generalizability. We improved the model performance with task voting and feature engineering and considered the generalizability by reserving a hold-out dataset. Secondly, these automatically extractable measures were noted to differ across languages and datasets^23,46^. Future studies should aim to develop a model on cross-lingual datasets with more subjects.

## Data Availability

All data produced in the present study are available upon reasonable request to the authors

## Acknowledgements

We appreciate all the participants and their families for the time and effort to contribute to this study.

## References

1. Volkmer S, Meyer-Lindenberg A, Schwarz E. Large language models in psychiatry: Opportunities and challenges. Psychiatry Research. 2024;339:116026. doi:10.1016/j.psychres.2024.116026

2. Low DM, Bentley KH, Ghosh SS. Automated assessment of psychiatric disorders using speech: A systematic review. Laryngoscope Investig Otolaryngol. 2020;5(1):96–116. doi:10.1002/lio2.354

3. He R, Palominos C, Zhang H, Alonso-Sánchez MF, Palaniyappan L, Hinzen W. Navigating the semantic space: Unraveling the structure of meaning in psychosis using different computational language models. Psychiatry Research. 2024;333:115752. doi:10.1016/j.psychres.2024.115752

4. Kliper R, Portuguese S, Weinshall D. Prosodic Analysis of Speech and the Underlying Mental State. In: Serino S, Matic A, Giakoumis D, Lopez G, Cipresso P, eds. Pervasive Computing Paradigms for Mental Health. Cham: Springer International Publishing; 2016:52–62. doi:10.1007/978-3-319-32270-4_6

5. Martínez-Sánchez F, Muela-Martínez JA, Cortés-Soto P, et al. Can the Acoustic Analysis of Expressive Prosody Discriminate Schizophrenia? Span J Psychol. 2015;18:E86. doi:10.1017/sjp.2015.85

6. Agurto C, Pietrowicz M, Norel R, et al. Analyzing acoustic and prosodic fluctuations in free speech to predict psychosis onset in high-risk youths. In: 2020 42nd Annual International Conference of the IEEE Engineering in Medicine & Biology Society (EMBC). ; 2020:5575–5579. doi:10.1109/EMBC44109.2020.9176841

7. Corcoran CM, Carrillo F, Fernández-Slezak D, et al. Prediction of psychosis across protocols and risk cohorts using automated language analysis. World Psychiatry. 2018;17(1):67–75. doi:10.1002/wps.20491

8. Rezaii N, Walker E, Wolff P. A machine learning approach to predicting psychosis using semantic density and latent content analysis. npj Schizophr. 2019;5(1):1–12. doi:10.1038/s41537-019-0077-9

9. Bianciardi B, Gajwani R, Gross J, et al. Investigating temporal and prosodic markers in clinical high-risk for psychosis participants using automated acoustic analysis. Early Interv Psychiatry. 2023;17(3):327–330. doi:10.1111/eip.13357

10. Figueroa-Barra A, Del Aguila D, Cerda M, et al. Automatic language analysis identifies and predicts schizophrenia in first-episode of psychosis. Schizophrenia (Heidelb). 2022;8(1):53. doi:10.1038/s41537-022-00259-3

11. Tang SX, Spilka MJ, John M, et al. Automated, Objective Speech and Language Markers of Longitudinal Changes in Psychosis Symptoms. July 2024:2024.07.19.24310718. doi:10.1101/2024.07.19.24310718

12. Morgan SE, Diederen K, Vértes PE, et al. Natural Language Processing markers in first episode psychosis and people at clinical high-risk. Transl Psychiatry. 2021;11(1):1–9. doi:10.1038/s41398-021-01722-y

13. Ben Moshe T, Ziv I, Dershowitz N, Bar K. The contribution of prosody to machine classification of schizophrenia. Schizophr. 2024;10(1):1–9. doi:10.1038/s41537-024-00463-3

14. Huang YJ, Lin YT, Liu CC, et al. Assessing Schizophrenia Patients Through Linguistic and Acoustic Features Using Deep Learning Techniques. IEEE Trans Neural Syst Rehabil Eng. 2022;30:947–956. doi:10.1109/TNSRE.2022.3163777

15. Voppel AE, de Boer JN, Brederoo SG, Schnack HG, Sommer IEC. Semantic and Acoustic Markers in Schizophrenia-Spectrum Disorders: A Combinatory Machine Learning Approach. Schizophrenia Bulletin. 2023;49(Supplement_2):S163–S171. doi:10.1093/schbul/sbac142

16. Radford A, Kim JW, Xu T, Brockman G, McLeavey C, Sutskever I. Robust Speech Recognition via Large-Scale Weak Supervision. December 2022. doi:10.48550/arXiv.2212.04356

17. Eyben F, Scherer KR, Schuller BW, et al. The Geneva Minimalistic Acoustic Parameter Set (GeMAPS) for Voice Research and Affective Computing. IEEE Trans Affective Comput. 2016;7(2):190–202. doi:10.1109/TAFFC.2015.2457417

18. Mertens P. The prosogram: semi-automatic transcription of prosody based on a tonal perception model. In: Proceedings of Speech Prosody 2004. Nara (Japan); 2004.

19. Ciampelli S, de Boer JN, Voppel AE, et al. Syntactic Network Analysis in Schizophrenia-Spectrum Disorders. Schizophrenia Bulletin. 2023;49(Supplement_2):S172-S182. doi:10.1093/schbul/sbac194

20. Ferrer-i-Cancho R, Gómez-Rodríguez C, Esteban JL, Alemany-Puig L. Optimality of syntactic dependency distances. Phys Rev E. 2022;105(1):014308. doi:10.1103/PhysRevE.105.014308

21. Ferrer-i-Cancho R, Gómez-Rodríguez C. Dependency distance minimization predicts compression. In: Čech R, Chen X, eds. Proceedings of the Second Workshop on Quantitative Syntax (Quasy, SyntaxFest 2021). Sofia, Bulgaria: Association for Computational Linguistics; 2021:45–57. https://aclanthology.org/2021.quasy-1.4. Accessed June 6, 2024.

22. de Boer JN, Brederoo SG, Voppel AE, Sommer IEC. Anomalies in language as a biomarker for schizophrenia. Current Opinion in Psychiatry. 2020;33(3):212. doi:10.1097/YCO.0000000000000595

23. Parola A, Lin JM, Simonsen A, et al. Speech disturbances in schizophrenia: Assessing cross-linguistic generalizability of NLP automated measures of coherence. Schizophrenia Research. August 2022. doi:10.1016/j.schres.2022.07.002

24. Grave E, Bojanowski P, Gupta P, Joulin A, Mikolov T. Learning Word Vectors for 157 Languages. In: Proceedings of the Eleventh International Conference on Language Resources and Evaluation (LREC 2018).; 2018. https://infoscience.epfl.ch/record/253313.

25. Gutiérrez-Fandiño A, Armengol-Estapé J, Pàmies M, et al. MarIA: Spanish Language Models. Procesamiento del Lenguaje Natural. 2022;68(0):39–60. http://journal.sepln.org/sepln/ojs/ojs/index.php/pln/article/view/6405. Accessed August 1, 2022.

26. Xu W, Portanova J, Chander A, Ben-Zeev D, Cohen T. The Centroid Cannot Hold: Comparing Sequential and Global Estimates of Coherence as Indicators of Formal Thought Disorder. AMIA Annu Symp Proc. 2020;2020:1315–1324.

27. Radford A, Kim JW, Hallacy C, et al. Learning Transferable Visual Models From Natural Language Supervision. February 2021. doi:10.48550/arXiv.2103.00020

28. Hayton JC, Allen DG, Scarpello V. Factor Retention Decisions in Exploratory Factor Analysis: a Tutorial on Parallel Analysis. Organizational Research Methods. 2004;7(2):191–205. doi:10.1177/1094428104263675

29. Corcoran CM, Mittal VA, Bearden CE, et al. Language as a biomarker for psychosis: A natural language processing approach. Schizophrenia Research. 2020;226:158–166. doi:10.1016/j.schres.2020.04.032

30. Hinzen W, Palaniyappan L. The ‘L-factor’: Language as a transdiagnostic dimension in psychopathology. Progress in Neuro-Psychopharmacology and Biological Psychiatry. 2024;131:110952. doi:10.1016/j.pnpbp.2024.110952

31. Schneider K, Leinweber K, Jamalabadi H, et al. Syntactic complexity and diversity of spontaneous speech production in schizophrenia spectrum and major depressive disorders. Schizophr. 2023;9(1):1–10. doi:10.1038/s41537-023-00359-8

32. Alonso-Sánchez MF, Ford SD, MacKinley M, Silva A, Limongi R, Palaniyappan L. Progressive changes in descriptive discourse in First Episode Schizophrenia: a longitudinal computational semantics study. Schizophrenia (Heidelb). 2022;8(1):36. doi:10.1038/s41537-022-00246-8

33. Boer JN, Voppel AE, Brederoo SG, et al. Acoustic speech markers for schizophrenia-spectrum disorders: a diagnostic and symptom-recognition tool. Psychological Medicine. 2023;53(4):1302–1312. doi:10.1017/S0033291721002804

34. Holmlund TB, Chandler C, Foltz PW, et al. Applying speech technologies to assess verbal memory in patients with serious mental illness. npj Digit Med. 2020;3(1):1–8. doi:10.1038/s41746-020-0241-7

35. Clarke N, Barrick TR, Garrard P. A Comparison of Connected Speech Tasks for Detecting Early Alzheimer’s Disease and Mild Cognitive Impairment Using Natural Language Processing and Machine Learning. Frontiers in Computer Science. 2021;3. 10.3389/fcomp.2021.634360. Accessed April 17, 2022.

36. Bose A, Dutta M, Dash NS, Nandi R, Dutt A, Ahmed S. Importance of Task Selection for Connected Speech Analysis in Patients with Alzheimer’s Disease from an Ethnically Diverse Sample. Journal of Alzheimer’s Disease. 2022;87(4):1475–1481. doi:10.3233/JAD-220166

37. Chen Yi, Hartsuiker RJ, Pistono A. A comparison of different connected-speech tasks for detecting mild cognitive impairment using multivariate pattern analysis. Aphasiology. May 2024:1–24. doi:10.1080/02687038.2024.2358556

38. Berardi M, Brosch K, Pfarr JK, et al. Relative importance of speech and voice features in the classification of schizophrenia and depression. Transl Psychiatry. 2023;13(1):1–8. doi:10.1038/s41398-023-02594-0

39. Tang SX, Kriz R, Cho S, et al. Natural language processing methods are sensitive to sub-clinical linguistic differences in schizophrenia spectrum disorders. NPJ Schizophr. 2021;7(1):25. doi:10.1038/s41537-021-00154-3

40. Fineberg SK, Deutsch-Link S, Ichinose M, et al. Word use in first-person accounts of schizophrenia. Br J Psychiatry. 2015;206(1):32–38. doi:10.1192/bjp.bp.113.140046

41. Lundin NB, Cowan HR, Singh DK, Moe AM. Lower cohesion and altered first-person pronoun usage in the spoken life narratives of individuals with schizophrenia. Schizophrenia Research. 2023;259:140–149. doi:10.1016/j.schres.2023.04.001

42. Cohen AS, Alpert M, Nienow TM, Dinzeo TJ, Docherty NM. Computerized measurement of negative symptoms in schizophrenia. J Psychiatr Res. 2008;42(10):827–836. doi:10.1016/j.jpsychires.2007.08.008

43. Mitra S, Mahintamani T, Kavoor AR, Nizamie SH. Negative symptoms in schizophrenia. Ind Psychiatry J. 2016;25(2):135–144. doi:10.4103/ipj.ipj_30_15

44. Stanislawski ER, Bilgrami ZR, Sarac C, et al. Negative symptoms and speech pauses in youths at clinical high risk for psychosis. npj Schizophr. 2021;7(1):1–3. doi:10.1038/s41537-020-00132-1

45. Zhao Q, Wang WQ, Fan HZ, et al. Vocal acoustic features may be objective biomarkers of negative symptoms in schizophrenia: A cross-sectional study. Schizophrenia Research. 2022;250:180–185. doi:10.1016/j.schres.2022.11.013

46. Stegmann GM, Hahn S, Liss J, et al. Repeatability of Commonly Used Speech and Language Features for Clinical Applications. Digit Biomark. 2020;4(3):109–122. doi:10.1159/000511671

