## supplementary information for "Task-voting for schizophrenia spectrum disorders prediction using machine learning across linguistic feature domains"

### 1. Prosogram features

Supplementary Table 1 listed the 31 Prosogram features used in this study and their definitions. These features were used together with the 88 eGeMAPs features.

Supplementary Table 1. Prosogram features with definitions.

| Variables | Definitions |
| --- | --- |
| SpeechRate | speech rate = NrOfNucl / (TotNuclDur + TotInternuclDur) |
| NrofSafe | number of nuclei without outliers and discontinuities |
| SpeechTime | speech time = (TotNuclDur + TotInternuclDur + TotPauseDur), for current speaker |
| TotNuclDur | total nucleus duration, for current speaker |
| TotInternuclDur | total internucleus duration, for current speaker |
| TotPauseDur | total pause duration, for current speaker |
| PropPause | TotPauseDur / (TotNuclDur + TotInternuclDur + TotPauseDur), for current speaker |
| F0MedianInST | median in ST of F0 values in Hz |
| PitchMeanST | mean in ST of F0 values in ST, 2 per nucleus: low and high |
| PitchStdevST | standard deviation of pitch values in ST, 2 per nucleus: low and high |
| PitchRange | pitch range (span), in ST |
| PitchTopST | top of pitch range, in ST |
| PitchBottomST | bottom of pitch range, in ST |
| RawF0_p25 | 25 percentile of raw F0 values in nuclei |
| RawF0_p50 | 50 percentile of raw F0 values in nuclei |
| RawF0_p75 | 75 percentile of raw F0 values in nuclei |
| RawF0_mean | mean of raw F0 values in nuclei |
| PropLevel | Proportion of nuclei without glissando, i.e. with level pitch, after stylization |
| Gliss | Proportion of nuclei with abs pitch change >= 4 ST |
| Rises | Proportion of nuclei with pitch change >= 4 ST |
| Falls | Proportion of nuclei with pitch change <= -4 ST |
| TrajIntra | Time-normalized pitch trajectory of intrasyllabic variations |
| TrajInter | Time-normalized pitch trajectory of intersyllabic variations |
| TrajPhon | Time-normalized pitch trajectory of all pitch variations |
| TrajIntraZ | Pitch range normalized TrajIntra |
| TrajInterZ | Pitch range normalized TrajInter |
| TrajPhonZ | Pitch range normalized TrajPhon |
| NuclDurMean | mean nucleus duration |
| NuclDurStdev | standard deviation of nucleus duration |
| NuclDurMedian | median nucleus duration |
| nPVI_nucldur | nPVI of nucleus duration |

### 2. Morphosyntactic features

Supplementary Table 2 listed the 76 morphosyntactic features used in this study and their definitions.

Supplementary Table 2. Morphosyntactic features with definitions.

| Variables | Definitions |
| --- | --- |
| TokenNum | Number of words without stopwords |
| WordNum | Number of words including stopwords |
| Sent_Num | Number of sentences |
| NP_Num | Number of noun chuncks (extracted from spaCy) |
| WordTTR | Type-token-ratio of words |
| stpw_ratio | Number of stopwords / Number of words |
| ADJ_PosR | Number of adjectives / Number of words |
| ADP_PosR | Number of adpositions / Number of words |
| ADV_PosR | Number of adverbs / Number of words |
| AUX_PosR | Number of auxilary verbs/ Number of words |
| CCONJ_PosR | Number of coordinating conjunctions / Number of words |
| DET_PosR | Number of determiners / Number of words |
| INTJ_PosR | Number of interjections / Number of words |
| NOUN_PosR | Number of nouns / Number of words |
| NUM_PosR | Number of numerals / Number of words |
| PART_PosR | Number of particles / Number of words |
| PRON_PosR | Number of pronouns / Number of words |
| PROPN_PosR | Number of proper nouns / Number of words |
| PUNCT_PosR | Number of punctuations / Number of words |
| SCONJ_PosR | Number of subordinating conjunctions / Number of words |
| SYM_PosR | Number of symbols / Number of words |
| VERB_PosR | Number of verbs / Number of words |
| acl_DepR | Number of acl (adjectival clause) dependency relations / Number of all dependency relations |
| advcl_DepR | Number of advcl (adverbial clause) dependency relations / Number of all dependency relations |
| advmod_DepR | Number of advmod (adverbial modifier) dependency relations / Number of all dependency relations |
| amod_DepR | Number of amod (adjectival modifier) dependency relations / Number of all dependency relations |
| appos_DepR | Number of appos (appositional modifier) dependency relations / Number of all dependency relations |
| aux_DepR | Number of aux (auxiliary) dependency relations / Number of all dependency relations |
| case_DepR | Number of case (case marking) dependency relations / Number of all dependency relations |
| cc_DepR | Number of cc (coordinating conjunction) dependency relations / Number of all dependency relations |
| ccomp_DepR | Number of ccomp (clausal complement) dependency relations / Number of all dependency relations |
| compound_DepR | Number of compound dependency relations / Number of all dependency relations |
| conj_DepR | Number of conj (conjunct) dependency relations / Number of all dependency relations |
| cop_DepR | Number of cop (copula) dependency relations / Number of all dependency relations |
| csubj_DepR | Number of csubj (clausal subject) dependency relations / Number of all dependency relations |
| dep_DepR | Number of dep (unclassified dependent) dependency relations / Number of all dependency relations |
| det_DepR | Number of det (determiner) dependency relations / Number of all dependency relations |
| fixed_DepR | Number of fixed (fixed multiword expression) dependency relations / Number of all dependency relations |
| flat_DepR | Number of flat (flat multiword expression) dependency relations / Number of all dependency relations |
| iobj_DepR | Number of iobj (indirect object) dependency relations / Number of all dependency relations |
| mark_DepR | Number of mark (marker) dependency relations / Number of all dependency relations |
| nmod_DepR | Number of nmod (nominal modifier) dependency relations / Number of all dependency relations |
| nsubj_DepR | Number of nsubj (nominal subject) dependency relations / Number of all dependency relations |
| nummod_DepR | Number of nummod (numeric modifier) dependency relations / Number of all dependency relations |
| obj_DepR | Number of obj (object) dependency relations / Number of all dependency relations |
| obl_DepR | Number of obl (oblique nominal) dependency relations / Number of all dependency relations |
| parataxis_DepR | Number of parataxis (paratactic relation) dependency relations / Number of all dependency relations |
| xcomp_DepR | Number of xcomp (open clausal complement) dependency relations / Number of all dependency relations |
| Fut_TenseR | Number of future tense / Number of words |
| Imp_TenseR | Number of imperfect tense / Number of words |
| Past_TenseR | Number of past tense / Number of words |
| Pres_TenseR | Number of present tense / Number of words |
| Person1R | Number of first person / Number of person-related features |
| Person2R | Number of second person / Number of person-related features |
| Person3R | Number of third person / Number of person-related features |
| Negation | Number of negation / Number of words |
| Ind_MoodR | Number of indicative mood / Number of words |
| Imp_MoodR | Number of imperative mood / Number of words |
| Sub_MoodR | Number of subjunctive mood / Number of words |
| Cnd_MoodR | Number of conditional mood / Number of words |
| MascR | Number of masculine gender / Number of words |
| FemR | Number of feminine gender / Number of words |
| SingR | Number of singular number / Number of words |
| PlurR | Number of plural number / Number of words |
| DefiniteR | Number of definiteness / Number of words |
| IndefiniteR | Number of indefiniteness / Number of words |
| ADD | Averaged dependency distance |
| MDD | Maximum dependency distance |
| NodeNum | Number of nodes |
| PhraseRatio | Number of phrasal nodes / Number of leave nodes |
| Depth | Maximum syntactic depth across all words |
| DepthMean | Mean syntactic depth across all words |
| DepthNorm | Maximum syntactic depth across all words / Number of words |
| DepthApEn | Approximate entropy of the array of syntactic depth, indicating how predictablility of syntactic depth patterns |
| DepthSkew | Skewness of the array of syntactic depth, a proxy for left-branching |
| Linearization | Correlation between syntactic depth and linear order of words |

### 3. Feature selection

These features were removed due to high correlations with other features:

TotNuclDur, TotInternuclDur, PitchMeanST, RawF0_p50, RawF0_mean, TrajPhon, F0semitoneFrom27_5Hz_sma3nz_percentile50_0, F0semitoneFrom27_5Hz_sma3nz_percentile80_0, loudness_sma3_percentile50_0, loudness_sma3_percentile80_0, loudness_sma3_meanFallingSlope, spectralFlux_sma3_amean, F1amplitudeLogRelF0_sma3nz_stddevNorm, F2amplitudeLogRelF0_sma3nz_amean, F2amplitudeLogRelF0_sma3nz_stddevNorm, F3amplitudeLogRelF0_sma3nz_amean, mfcc2V_sma3nz_amean, mfcc4V_sma3nz_amean, hammarbergIndexUV_sma3nz_amean, FT_Amp, FT_Kurt, FT_stat_MeanK1, FT_stat_Amp, BERT_MeanK2, BERT_MeanG, BERT_Amp, BERT_stat_MeanK1, BERT_stat_Amp, BERT_stat_Kurt, BERT_cuml_MeanK1, BERT_cuml_WL, BERT_cuml_Valley, BERT_cuml_Amp, BERT_cuml_Skew, BERT_cuml_Kurt, BERT_cuml_ApEn, BERT_cuml_Acf1, Sent_stat_MeanK1, Sent_cuml_MeanK1, WordNum, NP_Num, conj_DepR, det_DepR, Person3R, Depth

Supplementary Table 3 showed the model performance with different number of features. The bold line indicated the best aggregated accuracy score.

Supplementary Table 3. Model accuracies with difference number of features.

| No. of features | Cross validation accuracy | Hold out accuracy | Aggregated accuracy |
| --- | --- | --- | --- |
| 1 | 0.728 | 0.580 | 0.088 |
| 2 | 0.716 | 0.531 | 0.067 |
| 3 | 0.704 | 0.605 | 0.133 |
| 4 | 0.716 | 0.654 | 0.222 |
| 5 | 0.728 | 0.741 | 1.469 |
| 6 | 0.704 | 0.630 | 0.180 |
| 7 | 0.704 | 0.691 | 1.395 |
| 8 | 0.741 | 0.667 | 0.190 |
| 9 | 0.679 | 0.667 | 1.346 |
| 10 | 0.716 | 0.691 | 1.407 |
| 11 | 0.741 | 0.704 | 1.444 |
| 12 | 0.679 | 0.691 | 1.370 |
| 13 | 0.716 | 0.790 | 0.203 |
| 14 | 0.716 | 0.778 | 0.242 |
| 15 | 0.704 | 0.790 | 0.173 |
| 16 | 0.704 | 0.704 | 1.407 |
| 17 | 0.704 | 0.691 | 1.395 |
| 18 | 0.728 | 0.679 | 1.407 |
| 19 | 0.691 | 0.778 | 0.170 |
| 20 | 0.753 | 0.741 | 1.494 |
| 21 | 0.704 | 0.728 | 1.432 |
| 22 | 0.691 | 0.728 | 1.420 |
| 23 | 0.753 | 0.728 | 1.481 |
| **24** | **0.815** | **0.840** | **1.654** |
| 25 | 0.741 | 0.765 | 1.506 |
| 26 | 0.741 | 0.704 | 1.444 |
| 27 | 0.642 | 0.716 | 0.183 |
| 28 | 0.753 | 0.778 | 1.531 |
| 29 | 0.691 | 0.667 | 1.358 |
| 30 | 0.815 | 0.753 | 0.254 |
| 31 | 0.728 | 0.741 | 1.469 |
| 32 | 0.741 | 0.778 | 1.519 |
| 33 | 0.753 | 0.790 | 1.543 |
| 34 | 0.691 | 0.753 | 0.234 |
| 35 | 0.765 | 0.827 | 0.258 |
| 36 | 0.654 | 0.753 | 0.143 |
| 37 | 0.778 | 0.778 | 1.556 |
| 38 | 0.654 | 0.864 | 0.072 |
| 39 | 0.704 | 0.716 | 1.420 |
| 40 | 0.778 | 0.704 | 0.200 |
| 41 | 0.753 | 0.667 | 0.164 |
| 42 | 0.704 | 0.827 | 0.124 |
| 43 | 0.765 | 0.704 | 0.238 |
| 44 | 0.778 | 0.741 | 1.519 |
| 45 | 0.815 | 0.704 | 0.137 |
| 46 | 0.741 | 0.753 | 1.494 |
| 47 | 0.741 | 0.691 | 1.432 |
| 48 | 0.741 | 0.654 | 0.161 |
| 49 | 0.741 | 0.790 | 1.531 |
| 50 | 0.728 | 0.741 | 1.469 |
| 51 | 0.753 | 0.654 | 0.143 |
| 52 | 0.728 | 0.741 | 1.469 |
| 53 | 0.753 | 0.716 | 1.469 |
| 54 | 0.741 | 0.728 | 1.469 |
| 55 | 0.753 | 0.790 | 1.543 |
| 56 | 0.704 | 0.704 | 1.407 |
| 57 | 0.741 | 0.617 | 0.110 |
| 58 | 0.691 | 0.630 | 0.214 |
| 59 | 0.716 | 0.741 | 1.457 |
| 60 | 0.778 | 0.679 | 0.147 |
| 61 | 0.704 | 0.704 | 1.407 |
| 62 | 0.728 | 0.642 | 0.159 |
| 63 | 0.790 | 0.716 | 0.203 |
| 64 | 0.741 | 0.679 | 0.230 |
| 65 | 0.704 | 0.765 | 0.238 |
| 66 | 0.778 | 0.753 | 1.531 |
| 67 | 0.778 | 0.679 | 0.148 |
| 68 | 0.716 | 0.741 | 1.457 |
| 69 | 0.691 | 0.691 | 1.383 |
| 70 | 0.741 | 0.679 | 0.230 |
| 71 | 0.667 | 0.753 | 0.164 |
| 72 | 0.691 | 0.704 | 1.395 |
| 73 | 0.753 | 0.741 | 1.494 |
| 74 | 0.691 | 0.667 | 1.358 |
| 75 | 0.741 | 0.778 | 1.519 |
| 76 | 0.741 | 0.790 | 1.531 |
| 77 | 0.741 | 0.716 | 1.457 |
| 78 | 0.741 | 0.728 | 1.469 |
| 79 | 0.691 | 0.691 | 1.383 |
| 80 | 0.765 | 0.642 | 0.114 |
| 81 | 0.728 | 0.741 | 1.469 |
| 82 | 0.728 | 0.691 | 1.420 |
| 83 | 0.667 | 0.790 | 0.118 |
| 84 | 0.691 | 0.654 | 1.346 |
| 85 | 0.765 | 0.654 | 0.128 |
| 86 | 0.778 | 0.765 | 1.543 |
| 87 | 0.716 | 0.617 | 0.135 |
| 88 | 0.741 | 0.679 | 0.230 |
| 89 | 0.704 | 0.728 | 1.432 |
| 90 | 0.704 | 0.654 | 1.358 |
| 91 | 0.691 | 0.642 | 1.333 |
| 92 | 0.790 | 0.630 | 0.088 |
| 93 | 0.765 | 0.728 | 1.494 |
| 94 | 0.741 | 0.704 | 1.444 |
| 95 | 0.704 | 0.778 | 0.200 |
| 96 | 0.765 | 0.765 | 1.531 |
| 97 | 0.765 | 0.691 | 0.197 |
| 98 | 0.679 | 0.691 | 1.370 |
| 99 | 0.704 | 0.753 | 1.457 |
| 100 | 0.753 | 0.741 | 1.494 |
| 101 | 0.716 | 0.765 | 1.481 |
| 102 | 0.753 | 0.741 | 1.494 |
| 103 | 0.741 | 0.679 | 0.230 |
| 104 | 0.704 | 0.716 | 1.420 |
| 105 | 0.704 | 0.765 | 0.238 |
| 106 | 0.741 | 0.716 | 1.457 |
| 107 | 0.728 | 0.667 | 0.226 |
| 108 | 0.765 | 0.716 | 1.481 |
| 109 | 0.716 | 0.790 | 0.203 |
| 110 | 0.778 | 0.815 | 1.593 |
| 111 | 0.704 | 0.765 | 0.238 |
| 112 | 0.728 | 0.654 | 0.187 |
| 113 | 0.654 | 0.741 | 0.161 |
| 114 | 0.728 | 0.778 | 1.506 |
| 115 | 0.753 | 0.642 | 0.126 |
| 116 | 0.704 | 0.753 | 1.457 |
| 117 | 0.741 | 0.852 | 0.143 |
| 118 | 0.728 | 0.654 | 0.187 |
| 119 | 0.716 | 0.778 | 0.242 |
| 120 | 0.704 | 0.802 | 0.153 |
| 121 | 0.716 | 0.728 | 1.444 |
| 122 | 0.728 | 0.778 | 1.506 |
| 123 | 0.765 | 0.617 | 0.093 |
| 124 | 0.654 | 0.654 | 1.309 |
| 125 | 0.704 | 0.605 | 0.133 |
| 126 | 0.741 | 0.716 | 1.457 |
| 127 | 0.716 | 0.691 | 1.407 |
| 128 | 0.765 | 0.765 | 1.531 |
| 129 | 0.679 | 0.704 | 1.383 |
| 130 | 0.704 | 0.741 | 1.444 |
| 131 | 0.753 | 0.691 | 0.234 |
| 132 | 0.765 | 0.704 | 0.238 |
| 133 | 0.667 | 0.802 | 0.108 |
| 134 | 0.741 | 0.778 | 1.519 |
| 135 | 0.704 | 0.753 | 1.457 |
| 136 | 0.778 | 0.815 | 1.593 |
| 137 | 0.765 | 0.691 | 0.197 |
| 138 | 0.679 | 0.827 | 0.102 |
| 139 | 0.679 | 0.691 | 1.370 |
| 140 | 0.728 | 0.667 | 0.226 |
| 141 | 0.679 | 0.630 | 1.309 |
| 142 | 0.704 | 0.654 | 1.358 |
| 143 | 0.679 | 0.667 | 1.346 |
| 144 | 0.691 | 0.691 | 1.383 |
| 145 | 0.728 | 0.716 | 1.444 |
| 146 | 0.691 | 0.790 | 0.150 |
| 147 | 0.704 | 0.852 | 0.105 |
| 148 | 0.716 | 0.667 | 1.383 |
| 149 | 0.704 | 0.617 | 0.153 |
| 150 | 0.716 | 0.630 | 0.156 |
| 151 | 0.667 | 0.741 | 0.190 |
| 152 | 0.753 | 0.679 | 0.193 |
| 153 | 0.654 | 0.765 | 0.128 |
| 154 | 0.778 | 0.802 | 1.580 |
| 155 | 0.716 | 0.852 | 0.115 |
| 156 | 0.667 | 0.728 | 0.226 |
| 157 | 0.716 | 0.593 | 0.106 |
| 158 | 0.704 | 0.716 | 1.420 |
| 159 | 0.765 | 0.741 | 1.506 |
| 160 | 0.667 | 0.716 | 1.383 |
| 161 | 0.728 | 0.716 | 1.444 |
| 162 | 0.630 | 0.605 | 1.235 |
| 163 | 0.642 | 0.691 | 1.333 |
| 164 | 0.716 | 0.617 | 0.135 |
| 165 | 0.691 | 0.815 | 0.122 |
| 166 | 0.704 | 0.753 | 1.457 |
| 167 | 0.691 | 0.716 | 1.407 |
| 168 | 0.704 | 0.728 | 1.432 |
| 169 | 0.679 | 0.617 | 0.210 |
| 170 | 0.654 | 0.728 | 0.187 |
| 171 | 0.691 | 0.753 | 0.234 |
| 172 | 0.704 | 0.679 | 1.383 |
| 173 | 0.716 | 0.667 | 1.383 |
| 174 | 0.679 | 0.753 | 0.193 |
| 175 | 0.716 | 0.691 | 1.407 |
| 176 | 0.667 | 0.642 | 1.309 |
| 177 | 0.753 | 0.815 | 0.254 |
| 178 | 0.704 | 0.691 | 1.395 |
| 179 | 0.765 | 0.654 | 0.128 |
| 180 | 0.728 | 0.741 | 1.469 |
| 181 | 0.691 | 0.753 | 0.234 |
| 182 | 0.765 | 0.716 | 1.481 |
| 183 | 0.778 | 0.765 | 1.543 |
| 184 | 0.728 | 0.741 | 1.469 |
| 185 | 0.654 | 0.753 | 0.142 |
| 186 | 0.704 | 0.654 | 1.358 |
| 187 | 0.802 | 0.753 | 1.556 |
| 188 | 0.642 | 0.617 | 1.259 |
| 189 | 0.679 | 0.605 | 0.173 |
| 190 | 0.679 | 0.815 | 0.110 |
| 191 | 0.630 | 0.679 | 1.309 |
| 192 | 0.765 | 0.716 | 1.481 |
| 193 | 0.691 | 0.741 | 1.432 |
| 194 | 0.704 | 0.778 | 0.200 |
| 195 | 0.728 | 0.630 | 0.138 |
| 196 | 0.753 | 0.704 | 1.457 |
| 197 | 0.691 | 0.691 | 1.383 |
| 198 | 0.704 | 0.728 | 1.432 |
| 199 | 0.679 | 0.691 | 1.370 |
| 200 | 0.667 | 0.691 | 1.358 |
| 201 | 0.667 | 0.642 | 1.309 |
| 202 | 0.704 | 0.679 | 1.383 |
| 203 | 0.654 | 0.728 | 0.187 |
| 204 | 0.679 | 0.827 | 0.102 |
| 205 | 0.691 | 0.568 | 0.102 |
| 206 | 0.778 | 0.617 | 0.087 |
| 207 | 0.691 | 0.630 | 0.214 |
| 208 | 0.790 | 0.716 | 0.203 |
| 209 | 0.691 | 0.728 | 1.420 |
| 210 | 0.654 | 0.790 | 0.106 |
| 211 | 0.704 | 0.704 | 1.407 |
| 212 | 0.741 | 0.667 | 0.190 |
| 213 | 0.654 | 0.815 | 0.092 |
| 214 | 0.642 | 0.765 | 0.114 |
| 215 | 0.679 | 0.679 | 1.358 |
| 216 | 0.667 | 0.679 | 1.346 |
| 217 | 0.728 | 0.580 | 0.088 |
| 218 | 0.753 | 0.778 | 1.531 |
| 219 | 0.741 | 0.691 | 1.432 |
| 220 | 0.691 | 0.741 | 1.432 |
| 221 | 0.679 | 0.691 | 1.370 |
| 222 | 0.667 | 0.691 | 1.358 |
| 223 | 0.679 | 0.765 | 0.167 |
| 224 | 0.630 | 0.654 | 1.284 |
| 225 | 0.716 | 0.741 | 1.457 |
| 226 | 0.654 | 0.753 | 0.143 |
| 227 | 0.630 | 0.630 | 1.259 |
| 228 | 0.642 | 0.691 | 1.333 |
| 229 | 0.630 | 0.667 | 1.296 |
| 230 | 0.667 | 0.691 | 1.358 |
| 231 | 0.617 | 0.741 | 0.110 |
| 232 | 0.691 | 0.790 | 0.150 |
| 233 | 0.667 | 0.630 | 1.296 |
| 234 | 0.654 | 0.778 | 0.116 |
| 235 | 0.704 | 0.790 | 0.173 |
| 236 | 0.704 | 0.642 | 0.218 |
| 237 | 0.704 | 0.691 | 1.395 |
| 238 | 0.704 | 0.691 | 1.395 |
| 239 | 0.691 | 0.679 | 1.370 |
| 240 | 0.642 | 0.704 | 0.218 |
| 241 | 0.691 | 0.654 | 1.346 |
| 242 | 0.753 | 0.630 | 0.112 |
| 243 | 0.704 | 0.679 | 1.383 |
| 244 | 0.605 | 0.593 | 1.198 |
| 245 | 0.716 | 0.642 | 0.183 |
| 246 | 0.741 | 0.716 | 1.457 |
| 247 | 0.691 | 0.704 | 1.395 |
| 248 | 0.667 | 0.654 | 1.321 |
| 249 | 0.691 | 0.728 | 1.420 |
| 250 | 0.765 | 0.531 | 0.055 |
| 251 | 0.691 | 0.667 | 1.358 |
| 252 | 0.667 | 0.728 | 0.226 |
| 253 | 0.691 | 0.790 | 0.150 |
| 254 | 0.679 | 0.654 | 1.333 |
| 255 | 0.642 | 0.679 | 1.321 |
| 256 | 0.617 | 0.753 | 0.101 |
| 257 | 0.617 | 0.704 | 0.153 |
| 258 | 0.716 | 0.679 | 1.395 |
| 259 | 0.605 | 0.605 | 1.210 |
| 260 | 0.704 | 0.605 | 0.133 |
| 261 | 0.679 | 0.691 | 1.370 |
| 262 | 0.704 | 0.704 | 1.407 |
| 263 | 0.728 | 0.667 | 0.226 |
| 264 | 0.691 | 0.765 | 0.197 |
| 265 | 0.667 | 0.728 | 0.226 |
| 266 | 0.642 | 0.556 | 0.139 |
| 267 | 0.691 | 0.704 | 1.395 |
| 268 | 0.691 | 0.765 | 0.197 |
| 269 | 0.741 | 0.716 | 1.457 |
| 270 | 0.691 | 0.654 | 1.346 |
| 271 | 0.691 | 0.728 | 1.420 |
| 272 | 0.654 | 0.728 | 0.187 |
| 273 | 0.728 | 0.728 | 1.457 |
| 274 | 0.605 | 0.728 | 0.108 |

### 4. Exploratory factor analysis

Following the procedure in the guideline, we first performed a principal component analysis (PCA) on the real data to extract 23 components (Hayton et al., 2004). Then, we generated 100 random data matrices and fitted them with the same PCA models. The ratios of variance explained per component for the random data were averaged across these 100 instances. Only the first five factors explained more variance in the real data compared to the random data, as shown in Supplementary Figure 1.

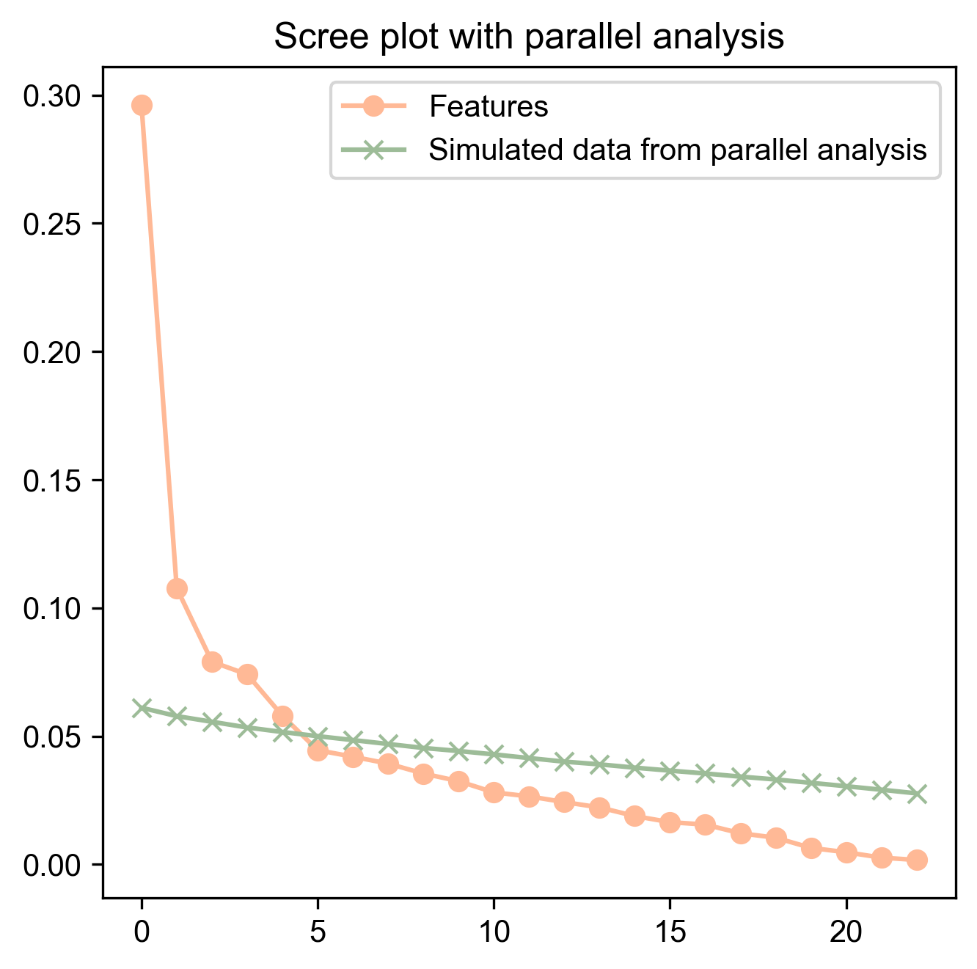

Supplementary Figure 1. Scree plot for PC-based parallel analysis.

*References*

Hayton, J. C., Allen, D. G., & Scarpello, V. (2004). Factor Retention Decisions in Exploratory Factor Analysis: A Tutorial on Parallel Analysis. *Organizational Research Methods*, *7*(2), 191–205. https://doi.org/10.1177/1094428104263675

### 5. Mixed-effected linear regression result summaries

Supplementary Table 4 summarized the mixed-effected linear regression results. *Z* values are the regression coefficients normalized by standard errors, reported with 95% confidence interval (CI). We also reported the uncorrected *p* values and corrected *q* values.

Supplementary Table 4. Regression results.

| Features | Z | *p* | *q* | 95% CI | |
| --- | --- | --- | --- | --- | --- |
|  |  |  |  | lower | upper |
| TrajPhonZ | -2.562 | 0.010 | 0.010 | -4.522 | -0.602 |
| loudness_sma3_stddevRisingSlope | -2.870 | 0.004 | 0.004 | -4.830 | -0.910 |
| FT_stat_Valley | 3.004 | 0.003 | 0.003 | 1.044 | 4.964 |
| Negation | 3.011 | 0.003 | 0.003 | 1.051 | 4.971 |
| shimmerLocaldB_sma3nz_amean | -3.017 | 0.003 | 0.003 | -4.977 | -1.057 |
| BERT_ApEn | -3.049 | 0.002 | 0.003 | -5.009 | -1.089 |
| DepthMean | -3.064 | 0.002 | 0.003 | -5.024 | -1.104 |
| PropLevel | -3.159 | 0.002 | 0.002 | -5.119 | -1.199 |
| BERT_stat_Valley | 3.284 | 0.001 | 0.002 | 1.324 | 5.244 |
| StddevUnvoicedSegmentLength | 3.295 | 0.001 | 0.002 | 1.335 | 5.255 |
| MeanVoicedSegmentLengthSec | 3.302 | 0.001 | 0.002 | 1.342 | 5.262 |
| TrajInter | -3.304 | 0.001 | 0.002 | -5.264 | -1.344 |
| pmt2rsp | -3.328 | 0.001 | 0.002 | -5.288 | -1.368 |
| PUNCT_PosR | 3.335 | 0.001 | 0.002 | 1.375 | 5.295 |
| Person1R | 3.448 | 0.001 | 0.001 | 1.488 | 5.408 |
| StddevVoicedSegmentLengthSec | 3.495 | 0.000 | 0.001 | 1.535 | 5.455 |
| nPVI_nucldur | 3.651 | 0.000 | 0.001 | 1.692 | 5.611 |
| mfcc3_sma3_amean | -3.851 | 0.000 | 0.000 | -5.811 | -1.891 |
| mfcc3V_sma3nz_amean | -3.980 | 0.000 | 0.000 | -5.940 | -2.021 |
| VoicedSegmentsPerSec | -4.044 | 0.000 | 0.000 | -6.004 | -2.084 |
| NuclDurStdev | 4.105 | 0.000 | 0.000 | 2.145 | 6.065 |
| NuclDurMedian | 5.091 | 0.000 | 0.000 | 3.131 | 7.051 |
| NuclDurMean | 5.163 | 0.000 | 0.000 | 3.203 | 7.123 |
| SpeechRate | -6.461 | 0.000 | 0.000 | -8.421 | -4.501 |

### 6. Correlation result summaries

Supplementary Table 5. Correlation results.

| Items | Features | r | CI95% | p | power | q |
| --- | --- | --- | --- | --- | --- | --- |
| P1 | SpeechRate | -0.192 | [-0.45 0.1 ] | 0.191 | 0.260 | 0.319 |
|  | NuclDurMean | 0.199 | [-0.09 0.46] | 0.174 | 0.277 | 0.435 |
|  | NuclDurMedian | 0.202 | [-0.09 0.46] | 0.169 | 0.283 | 0.281 |
|  | NuclDurStdev | 0.204 | [-0.08 0.46] | 0.164 | 0.289 | 0.717 |
|  | VoicedSegmentsPerSec | -0.277 | [-0.52 0.01] | 0.057 | 0.485 | 0.155 |
|  | mfcc3V_sma3nz_amean | -0.145 | [-0.41 0.15] | 0.326 | 0.167 | 0.651 |
|  | mfcc3_sma3_amean | -0.119 | [-0.39 0.17] | 0.421 | 0.127 | 0.843 |
|  | nPVI_nucldur | 0.237 | [-0.05 0.49] | 0.105 | 0.373 | 0.523 |
|  | StddevVoicedSegmentLengthSec | 0.249 | [-0.04 0.5 ] | 0.088 | 0.406 | 0.292 |
|  | Person1R | -0.216 | [-0.47 0.07] | 0.141 | 0.317 | 0.298 |
|  | PUNCT_PosR | 0.020 | [-0.27 0.3 ] | 0.893 | 0.052 | 0.904 |
|  | pmt2rsp | -0.524 | [-0.7 -0.28] | 0.000 | 0.976 | 0.001 |
|  | TrajInter | -0.022 | [-0.3 0.26] | 0.882 | 0.052 | 0.983 |
|  | MeanVoicedSegmentLengthSec | 0.173 | [-0.12 0.44] | 0.239 | 0.220 | 0.598 |
|  | StddevUnvoicedSegmentLength | 0.236 | [-0.05 0.49] | 0.107 | 0.369 | 0.267 |
|  | BERT_stat_Valley | 0.341 | [0.06 0.57] | 0.018 | 0.670 | 0.045 |
|  | PropLevel | -0.034 | [-0.31 0.25] | 0.820 | 0.056 | 0.995 |
|  | DepthMean | -0.216 | [-0.47 0.07] | 0.140 | 0.318 | 0.233 |
|  | BERT_ApEn | -0.193 | [-0.45 0.1 ] | 0.189 | 0.262 | 0.473 |
|  | shimmerLocaldB_sma3nz_amean | -0.198 | [-0.46 0.09] | 0.178 | 0.273 | 0.310 |
|  | Negation | 0.214 | [-0.07 0.47] | 0.144 | 0.313 | 0.760 |
|  | FT_stat_Valley | 0.196 | [-0.09 0.45] | 0.181 | 0.270 | 0.454 |
|  | loudness_sma3_stddevRisingSlope | -0.019 | [-0.3 0.27] | 0.897 | 0.052 | 0.904 |
|  | TrajPhonZ | -0.219 | [-0.47 0.07] | 0.135 | 0.326 | 0.737 |
| P2 | SpeechRate | -0.155 | [-0.42 0.13] | 0.292 | 0.185 | 0.365 |
|  | NuclDurMean | 0.133 | [-0.16 0.4 ] | 0.367 | 0.148 | 0.612 |
|  | NuclDurMedian | 0.234 | [-0.05 0.49] | 0.109 | 0.366 | 0.218 |
|  | NuclDurStdev | 0.005 | [-0.28 0.29] | 0.972 | 0.050 | 0.972 |
|  | VoicedSegmentsPerSec | -0.182 | [-0.44 0.11] | 0.215 | 0.239 | 0.430 |
|  | mfcc3V_sma3nz_amean | -0.156 | [-0.42 0.13] | 0.289 | 0.187 | 0.651 |
|  | mfcc3_sma3_amean | -0.137 | [-0.41 0.15] | 0.352 | 0.154 | 0.843 |
|  | nPVI_nucldur | 0.082 | [-0.21 0.36] | 0.578 | 0.086 | 0.971 |
|  | StddevVoicedSegmentLengthSec | -0.023 | [-0.31 0.26] | 0.874 | 0.053 | 0.969 |
|  | Person1R | -0.211 | [-0.47 0.08] | 0.149 | 0.306 | 0.298 |
|  | PUNCT_PosR | 0.111 | [-0.18 0.38] | 0.453 | 0.117 | 0.904 |
|  | pmt2rsp | -0.435 | [-0.64 -0.17] | 0.002 | 0.884 | 0.010 |
|  | TrajInter | 0.138 | [-0.15 0.41] | 0.348 | 0.156 | 0.871 |
|  | MeanVoicedSegmentLengthSec | 0.004 | [-0.28 0.29] | 0.979 | 0.050 | 0.979 |
|  | StddevUnvoicedSegmentLength | 0.181 | [-0.11 0.44] | 0.218 | 0.236 | 0.388 |
|  | BERT_stat_Valley | 0.223 | [-0.07 0.48] | 0.128 | 0.335 | 0.256 |
|  | PropLevel | -0.104 | [-0.38 0.19] | 0.482 | 0.108 | 0.789 |
|  | DepthMean | -0.110 | [-0.38 0.18] | 0.455 | 0.116 | 0.650 |
|  | BERT_ApEn | -0.059 | [-0.34 0.23] | 0.689 | 0.068 | 0.856 |
|  | shimmerLocaldB_sma3nz_amean | -0.312 | [-0.55 -0.03] | 0.031 | 0.589 | 0.309 |
|  | Negation | 0.019 | [-0.27 0.3 ] | 0.899 | 0.052 | 0.899 |
|  | FT_stat_Valley | 0.124 | [-0.17 0.39] | 0.402 | 0.134 | 0.523 |
|  | loudness_sma3_stddevRisingSlope | -0.018 | [-0.3 0.27] | 0.904 | 0.051 | 0.904 |
|  | TrajPhonZ | -0.001 | [-0.29 0.28] | 0.994 | 0.050 | 0.994 |
| P3 | SpeechRate | -0.159 | [-0.42 0.13] | 0.279 | 0.193 | 0.365 |
|  | NuclDurMean | 0.073 | [-0.22 0.35] | 0.621 | 0.078 | 0.792 |
|  | NuclDurMedian | 0.097 | [-0.19 0.37] | 0.513 | 0.100 | 0.641 |
|  | NuclDurStdev | 0.069 | [-0.22 0.35] | 0.639 | 0.075 | 0.972 |
|  | VoicedSegmentsPerSec | -0.144 | [-0.41 0.15] | 0.328 | 0.166 | 0.468 |
|  | mfcc3V_sma3nz_amean | -0.147 | [-0.41 0.14] | 0.317 | 0.171 | 0.651 |
|  | mfcc3_sma3_amean | -0.135 | [-0.4 0.16] | 0.361 | 0.150 | 0.843 |
|  | nPVI_nucldur | 0.059 | [-0.23 0.34] | 0.691 | 0.068 | 0.971 |
|  | StddevVoicedSegmentLengthSec | 0.158 | [-0.13 0.42] | 0.284 | 0.189 | 0.680 |
|  | Person1R | -0.158 | [-0.42 0.13] | 0.283 | 0.190 | 0.404 |
|  | PUNCT_PosR | 0.045 | [-0.24 0.32] | 0.763 | 0.060 | 0.904 |
|  | pmt2rsp | -0.305 | [-0.54 -0.02] | 0.035 | 0.567 | 0.088 |
|  | TrajInter | -0.041 | [-0.32 0.25] | 0.784 | 0.058 | 0.983 |
|  | MeanVoicedSegmentLengthSec | 0.083 | [-0.21 0.36] | 0.575 | 0.087 | 0.979 |
|  | StddevUnvoicedSegmentLength | 0.140 | [-0.15 0.41] | 0.342 | 0.159 | 0.453 |
|  | BERT_stat_Valley | 0.399 | [0.13 0.61] | 0.005 | 0.815 | 0.017 |
|  | PropLevel | 0.016 | [-0.27 0.3 ] | 0.913 | 0.051 | 0.995 |
|  | DepthMean | -0.281 | [-0.52 0. ] | 0.053 | 0.498 | 0.137 |
|  | BERT_ApEn | -0.205 | [-0.46 0.08] | 0.163 | 0.290 | 0.473 |
|  | shimmerLocaldB_sma3nz_amean | -0.194 | [-0.45 0.09] | 0.185 | 0.266 | 0.310 |
|  | Negation | 0.167 | [-0.12 0.43] | 0.257 | 0.207 | 0.760 |
|  | FT_stat_Valley | 0.223 | [-0.07 0.48] | 0.128 | 0.335 | 0.454 |
|  | loudness_sma3_stddevRisingSlope | -0.060 | [-0.34 0.23] | 0.684 | 0.069 | 0.865 |
|  | TrajPhonZ | -0.040 | [-0.32 0.25] | 0.790 | 0.058 | 0.959 |
| N1 | SpeechRate | -0.404 | [-0.62 -0.14] | 0.004 | 0.826 | 0.022 |
|  | NuclDurMean | 0.429 | [0.16 0.64] | 0.002 | 0.873 | 0.012 |
|  | NuclDurMedian | 0.541 | [0.3 0.72] | 0.000 | 0.984 | 0.001 |
|  | NuclDurStdev | 0.124 | [-0.17 0.39] | 0.400 | 0.135 | 0.972 |
|  | VoicedSegmentsPerSec | -0.615 | [-0.77 -0.4 ] | 0.000 | 0.998 | 0.000 |
|  | mfcc3V_sma3nz_amean | -0.027 | [-0.31 0.26] | 0.854 | 0.054 | 0.877 |
|  | mfcc3_sma3_amean | -0.136 | [-0.4 0.15] | 0.355 | 0.153 | 0.843 |
|  | nPVI_nucldur | 0.022 | [-0.26 0.3 ] | 0.882 | 0.052 | 0.971 |
|  | StddevVoicedSegmentLengthSec | 0.352 | [0.08 0.58] | 0.014 | 0.701 | 0.142 |
|  | Person1R | -0.297 | [-0.54 -0.01] | 0.041 | 0.544 | 0.257 |
|  | PUNCT_PosR | -0.070 | [-0.35 0.22] | 0.635 | 0.076 | 0.904 |
|  | pmt2rsp | -0.348 | [-0.58 -0.07] | 0.015 | 0.691 | 0.051 |
|  | TrajInter | -0.376 | [-0.6 -0.1] | 0.008 | 0.764 | 0.042 |
|  | MeanVoicedSegmentLengthSec | 0.512 | [0.27 0.7 ] | 0.000 | 0.969 | 0.002 |
|  | StddevUnvoicedSegmentLength | 0.481 | [0.23 0.67] | 0.001 | 0.944 | 0.005 |
|  | BERT_stat_Valley | 0.397 | [0.13 0.61] | 0.005 | 0.812 | 0.017 |
|  | PropLevel | -0.117 | [-0.39 0.17] | 0.429 | 0.125 | 0.789 |
|  | DepthMean | -0.297 | [-0.54 -0.01] | 0.040 | 0.546 | 0.137 |
|  | BERT_ApEn | -0.331 | [-0.56 -0.05] | 0.022 | 0.643 | 0.108 |
|  | shimmerLocaldB_sma3nz_amean | -0.140 | [-0.41 0.15] | 0.342 | 0.159 | 0.427 |
|  | Negation | 0.172 | [-0.12 0.43] | 0.242 | 0.217 | 0.760 |
|  | FT_stat_Valley | 0.206 | [-0.08 0.46] | 0.160 | 0.293 | 0.454 |
|  | loudness_sma3_stddevRisingSlope | -0.407 | [-0.62 -0.14] | 0.004 | 0.832 | 0.041 |
|  | TrajPhonZ | -0.026 | [-0.31 0.26] | 0.863 | 0.053 | 0.959 |
| N4 | SpeechRate | -0.209 | [-0.47 0.08] | 0.155 | 0.299 | 0.309 |
|  | NuclDurMean | 0.229 | [-0.06 0.48] | 0.118 | 0.351 | 0.392 |
|  | NuclDurMedian | 0.251 | [-0.04 0.5 ] | 0.086 | 0.410 | 0.214 |
|  | NuclDurStdev | 0.182 | [-0.11 0.44] | 0.215 | 0.238 | 0.717 |
|  | VoicedSegmentsPerSec | -0.271 | [-0.52 0.01] | 0.062 | 0.470 | 0.155 |
|  | mfcc3V_sma3nz_amean | -0.026 | [-0.31 0.26] | 0.862 | 0.053 | 0.877 |
|  | mfcc3_sma3_amean | 0.022 | [-0.26 0.3 ] | 0.881 | 0.052 | 0.922 |
|  | nPVI_nucldur | 0.027 | [-0.26 0.31] | 0.855 | 0.054 | 0.971 |
|  | StddevVoicedSegmentLengthSec | 0.141 | [-0.15 0.41] | 0.340 | 0.160 | 0.680 |
|  | Person1R | -0.189 | [-0.45 0.1 ] | 0.198 | 0.254 | 0.329 |
|  | PUNCT_PosR | -0.237 | [-0.49 0.05] | 0.105 | 0.372 | 0.886 |
|  | pmt2rsp | -0.158 | [-0.42 0.13] | 0.285 | 0.189 | 0.285 |
|  | TrajInter | -0.066 | [-0.34 0.22] | 0.655 | 0.073 | 0.983 |
|  | MeanVoicedSegmentLengthSec | 0.206 | [-0.08 0.46] | 0.159 | 0.294 | 0.530 |
|  | StddevUnvoicedSegmentLength | 0.176 | [-0.11 0.44] | 0.233 | 0.224 | 0.388 |
|  | BERT_stat_Valley | 0.130 | [-0.16 0.4 ] | 0.379 | 0.143 | 0.473 |
|  | PropLevel | -0.214 | [-0.47 0.07] | 0.144 | 0.313 | 0.719 |
|  | DepthMean | -0.063 | [-0.34 0.23] | 0.672 | 0.071 | 0.672 |
|  | BERT_ApEn | -0.029 | [-0.31 0.26] | 0.847 | 0.054 | 0.856 |
|  | shimmerLocaldB_sma3nz_amean | -0.218 | [-0.47 0.07] | 0.136 | 0.324 | 0.310 |
|  | Negation | 0.123 | [-0.17 0.39] | 0.405 | 0.133 | 0.760 |
|  | FT_stat_Valley | 0.028 | [-0.26 0.31] | 0.849 | 0.054 | 0.849 |
|  | loudness_sma3_stddevRisingSlope | -0.091 | [-0.37 0.2 ] | 0.539 | 0.094 | 0.865 |
|  | TrajPhonZ | 0.047 | [-0.24 0.33] | 0.753 | 0.061 | 0.959 |
| N6 | SpeechRate | -0.527 | [-0.71 -0.29] | 0.000 | 0.977 | 0.001 |
|  | NuclDurMean | 0.454 | [0.19 0.65] | 0.001 | 0.912 | 0.012 |
|  | NuclDurMedian | 0.484 | [0.23 0.68] | 0.000 | 0.947 | 0.002 |
|  | NuclDurStdev | 0.226 | [-0.06 0.48] | 0.123 | 0.343 | 0.717 |
|  | VoicedSegmentsPerSec | -0.428 | [-0.64 -0.16] | 0.002 | 0.873 | 0.012 |
|  | mfcc3V_sma3nz_amean | 0.095 | [-0.19 0.37] | 0.520 | 0.099 | 0.866 |
|  | mfcc3_sma3_amean | -0.027 | [-0.31 0.26] | 0.856 | 0.053 | 0.922 |
|  | nPVI_nucldur | 0.252 | [-0.03 0.5 ] | 0.084 | 0.414 | 0.523 |
|  | StddevVoicedSegmentLengthSec | 0.259 | [-0.03 0.51] | 0.076 | 0.433 | 0.292 |
|  | Person1R | -0.267 | [-0.51 0.02] | 0.067 | 0.456 | 0.257 |
|  | PUNCT_PosR | -0.023 | [-0.31 0.26] | 0.874 | 0.053 | 0.904 |
|  | pmt2rsp | -0.259 | [-0.51 0.03] | 0.076 | 0.434 | 0.094 |
|  | TrajInter | -0.392 | [-0.61 -0.12] | 0.006 | 0.801 | 0.042 |
|  | MeanVoicedSegmentLengthSec | 0.290 | [0.01 0.53] | 0.046 | 0.523 | 0.230 |
|  | StddevUnvoicedSegmentLength | 0.377 | [0.1 0.6] | 0.008 | 0.766 | 0.041 |
|  | BERT_stat_Valley | 0.436 | [0.17 0.64] | 0.002 | 0.885 | 0.017 |
|  | PropLevel | -0.236 | [-0.49 0.05] | 0.106 | 0.371 | 0.719 |
|  | DepthMean | -0.331 | [-0.56 -0.05] | 0.022 | 0.642 | 0.137 |
|  | BERT_ApEn | -0.396 | [-0.61 -0.13] | 0.005 | 0.809 | 0.054 |
|  | shimmerLocaldB_sma3nz_amean | -0.194 | [-0.45 0.1 ] | 0.186 | 0.265 | 0.310 |
|  | Negation | 0.048 | [-0.24 0.33] | 0.745 | 0.062 | 0.899 |
|  | FT_stat_Valley | 0.377 | [0.1 0.6] | 0.008 | 0.765 | 0.083 |
|  | loudness_sma3_stddevRisingSlope | -0.368 | [-0.59 -0.09] | 0.010 | 0.744 | 0.050 |
|  | TrajPhonZ | -0.026 | [-0.31 0.26] | 0.863 | 0.053 | 0.959 |
| G2 | SpeechRate | 0.214 | [-0.07 0.47] | 0.144 | 0.312 | 0.309 |
|  | NuclDurMean | -0.175 | [-0.44 0.11] | 0.233 | 0.224 | 0.467 |
|  | NuclDurMedian | -0.252 | [-0.5 0.03] | 0.083 | 0.416 | 0.214 |
|  | NuclDurStdev | -0.034 | [-0.32 0.25] | 0.819 | 0.056 | 0.972 |
|  | VoicedSegmentsPerSec | 0.027 | [-0.26 0.31] | 0.853 | 0.054 | 0.853 |
|  | mfcc3V_sma3nz_amean | -0.252 | [-0.5 0.04] | 0.085 | 0.413 | 0.423 |
|  | mfcc3_sma3_amean | -0.088 | [-0.36 0.2 ] | 0.554 | 0.091 | 0.849 |
|  | nPVI_nucldur | -0.036 | [-0.32 0.25] | 0.808 | 0.057 | 0.971 |
|  | StddevVoicedSegmentLengthSec | -0.057 | [-0.34 0.23] | 0.698 | 0.067 | 0.969 |
|  | Person1R | -0.067 | [-0.34 0.22] | 0.653 | 0.073 | 0.653 |
|  | PUNCT_PosR | 0.198 | [-0.09 0.46] | 0.177 | 0.274 | 0.886 |
|  | pmt2rsp | -0.264 | [-0.51 0.02] | 0.069 | 0.450 | 0.094 |
|  | TrajInter | 0.246 | [-0.04 0.5 ] | 0.091 | 0.398 | 0.305 |
|  | MeanVoicedSegmentLengthSec | -0.052 | [-0.33 0.24] | 0.724 | 0.064 | 0.979 |
|  | StddevUnvoicedSegmentLength | -0.026 | [-0.31 0.26] | 0.862 | 0.053 | 0.862 |
|  | BERT_stat_Valley | -0.043 | [-0.32 0.24] | 0.772 | 0.059 | 0.858 |
|  | PropLevel | 0.108 | [-0.18 0.38] | 0.464 | 0.114 | 0.789 |
|  | DepthMean | -0.277 | [-0.52 0.01] | 0.056 | 0.487 | 0.137 |
|  | BERT_ApEn | 0.027 | [-0.26 0.31] | 0.856 | 0.053 | 0.856 |
|  | shimmerLocaldB_sma3nz_amean | -0.019 | [-0.3 0.27] | 0.900 | 0.051 | 0.900 |
|  | Negation | -0.085 | [-0.36 0.2 ] | 0.565 | 0.089 | 0.808 |
|  | FT_stat_Valley | -0.158 | [-0.42 0.13] | 0.285 | 0.189 | 0.523 |
|  | loudness_sma3_stddevRisingSlope | 0.157 | [-0.13 0.42] | 0.288 | 0.187 | 0.575 |
|  | TrajPhonZ | 0.212 | [-0.08 0.47] | 0.147 | 0.308 | 0.737 |
| G5 | SpeechRate | -0.232 | [-0.48 0.06] | 0.112 | 0.360 | 0.309 |
|  | NuclDurMean | 0.071 | [-0.22 0.35] | 0.634 | 0.076 | 0.792 |
|  | NuclDurMedian | 0.134 | [-0.16 0.4 ] | 0.363 | 0.150 | 0.518 |
|  | NuclDurStdev | -0.014 | [-0.3 0.27] | 0.926 | 0.051 | 0.972 |
|  | VoicedSegmentsPerSec | -0.052 | [-0.33 0.24] | 0.724 | 0.064 | 0.804 |
|  | mfcc3V_sma3nz_amean | 0.023 | [-0.26 0.31] | 0.877 | 0.052 | 0.877 |
|  | mfcc3_sma3_amean | 0.079 | [-0.21 0.36] | 0.594 | 0.083 | 0.849 |
|  | nPVI_nucldur | 0.005 | [-0.28 0.29] | 0.971 | 0.050 | 0.971 |
|  | StddevVoicedSegmentLengthSec | 0.006 | [-0.28 0.29] | 0.969 | 0.050 | 0.969 |
|  | Person1R | -0.097 | [-0.37 0.19] | 0.513 | 0.100 | 0.570 |
|  | PUNCT_PosR | 0.105 | [-0.18 0.38] | 0.476 | 0.110 | 0.904 |
|  | pmt2rsp | -0.203 | [-0.46 0.09] | 0.166 | 0.286 | 0.184 |
|  | TrajInter | -0.003 | [-0.29 0.28] | 0.983 | 0.050 | 0.983 |
|  | MeanVoicedSegmentLengthSec | -0.008 | [-0.29 0.28] | 0.955 | 0.050 | 0.979 |
|  | StddevUnvoicedSegmentLength | 0.041 | [-0.25 0.32] | 0.784 | 0.058 | 0.862 |
|  | BERT_stat_Valley | 0.015 | [-0.27 0.3 ] | 0.917 | 0.051 | 0.917 |
|  | PropLevel | -0.088 | [-0.36 0.2 ] | 0.551 | 0.092 | 0.789 |
|  | DepthMean | -0.079 | [-0.36 0.21] | 0.594 | 0.083 | 0.672 |
|  | BERT_ApEn | -0.077 | [-0.35 0.21] | 0.603 | 0.081 | 0.856 |
|  | shimmerLocaldB_sma3nz_amean | -0.073 | [-0.35 0.22] | 0.624 | 0.078 | 0.694 |
|  | Negation | -0.110 | [-0.38 0.18] | 0.456 | 0.116 | 0.760 |
|  | FT_stat_Valley | 0.120 | [-0.17 0.39] | 0.418 | 0.128 | 0.523 |
|  | loudness_sma3_stddevRisingSlope | -0.158 | [-0.42 0.13] | 0.282 | 0.191 | 0.575 |
|  | TrajPhonZ | 0.109 | [-0.18 0.38] | 0.462 | 0.114 | 0.959 |
| G6 | SpeechRate | 0.031 | [-0.25 0.31] | 0.832 | 0.055 | 0.846 |
|  | NuclDurMean | 0.022 | [-0.26 0.3 ] | 0.880 | 0.052 | 0.880 |
|  | NuclDurMedian | 0.018 | [-0.27 0.3 ] | 0.901 | 0.051 | 0.974 |
|  | NuclDurStdev | 0.042 | [-0.25 0.32] | 0.777 | 0.059 | 0.972 |
|  | VoicedSegmentsPerSec | -0.147 | [-0.41 0.14] | 0.320 | 0.170 | 0.468 |
|  | mfcc3V_sma3nz_amean | -0.295 | [-0.53 -0.01] | 0.041 | 0.540 | 0.415 |
|  | mfcc3_sma3_amean | -0.185 | [-0.45 0.1 ] | 0.209 | 0.244 | 0.843 |
|  | nPVI_nucldur | 0.163 | [-0.13 0.43] | 0.269 | 0.199 | 0.897 |
|  | StddevVoicedSegmentLengthSec | 0.040 | [-0.25 0.32] | 0.787 | 0.058 | 0.969 |
|  | Person1R | -0.119 | [-0.39 0.17] | 0.420 | 0.128 | 0.525 |
|  | PUNCT_PosR | -0.018 | [-0.3 0.27] | 0.904 | 0.051 | 0.904 |
|  | pmt2rsp | -0.273 | [-0.52 0.01] | 0.060 | 0.476 | 0.094 |
|  | TrajInter | 0.006 | [-0.28 0.29] | 0.969 | 0.050 | 0.983 |
|  | MeanVoicedSegmentLengthSec | 0.019 | [-0.27 0.3 ] | 0.897 | 0.052 | 0.979 |
|  | StddevUnvoicedSegmentLength | 0.248 | [-0.04 0.5 ] | 0.090 | 0.402 | 0.267 |
|  | BERT_stat_Valley | 0.163 | [-0.13 0.43] | 0.268 | 0.199 | 0.447 |
|  | PropLevel | 0.001 | [-0.28 0.28] | 0.995 | 0.050 | 0.995 |
|  | DepthMean | -0.265 | [-0.51 0.02] | 0.069 | 0.451 | 0.137 |
|  | BERT_ApEn | -0.071 | [-0.35 0.22] | 0.630 | 0.077 | 0.856 |
|  | shimmerLocaldB_sma3nz_amean | -0.148 | [-0.41 0.14] | 0.316 | 0.172 | 0.427 |
|  | Negation | -0.127 | [-0.4 0.16] | 0.389 | 0.139 | 0.760 |
|  | FT_stat_Valley | 0.028 | [-0.26 0.31] | 0.848 | 0.054 | 0.849 |
|  | loudness_sma3_stddevRisingSlope | -0.059 | [-0.34 0.23] | 0.692 | 0.068 | 0.865 |
|  | TrajPhonZ | 0.066 | [-0.22 0.34] | 0.655 | 0.073 | 0.959 |
| G9 | SpeechRate | -0.029 | [-0.31 0.26] | 0.846 | 0.054 | 0.846 |
|  | NuclDurMean | -0.031 | [-0.31 0.25] | 0.832 | 0.055 | 0.880 |
|  | NuclDurMedian | 0.005 | [-0.28 0.29] | 0.974 | 0.050 | 0.974 |
|  | NuclDurStdev | -0.065 | [-0.34 0.22] | 0.661 | 0.072 | 0.972 |
|  | VoicedSegmentsPerSec | 0.095 | [-0.19 0.37] | 0.521 | 0.098 | 0.652 |
|  | mfcc3V_sma3nz_amean | -0.071 | [-0.35 0.22] | 0.632 | 0.077 | 0.877 |
|  | mfcc3_sma3_amean | 0.015 | [-0.27 0.3 ] | 0.922 | 0.051 | 0.922 |
|  | nPVI_nucldur | 0.069 | [-0.22 0.35] | 0.641 | 0.075 | 0.971 |
|  | StddevVoicedSegmentLengthSec | 0.020 | [-0.27 0.3 ] | 0.892 | 0.052 | 0.969 |
|  | Person1R | -0.258 | [-0.5 0.03] | 0.077 | 0.430 | 0.257 |
|  | PUNCT_PosR | -0.147 | [-0.41 0.14] | 0.318 | 0.171 | 0.904 |
|  | pmt2rsp | -0.292 | [-0.53 -0.01] | 0.044 | 0.531 | 0.088 |
|  | TrajInter | 0.098 | [-0.19 0.37] | 0.506 | 0.102 | 0.983 |
|  | MeanVoicedSegmentLengthSec | -0.073 | [-0.35 0.22] | 0.623 | 0.078 | 0.979 |
|  | StddevUnvoicedSegmentLength | -0.134 | [-0.4 0.16] | 0.362 | 0.150 | 0.453 |
|  | BERT_stat_Valley | 0.139 | [-0.15 0.41] | 0.345 | 0.157 | 0.473 |
|  | PropLevel | 0.088 | [-0.2 0.36] | 0.553 | 0.091 | 0.789 |
|  | DepthMean | 0.072 | [-0.22 0.35] | 0.626 | 0.078 | 0.672 |
|  | BERT_ApEn | 0.126 | [-0.16 0.4 ] | 0.394 | 0.137 | 0.789 |
|  | shimmerLocaldB_sma3nz_amean | -0.224 | [-0.48 0.06] | 0.126 | 0.338 | 0.310 |
|  | Negation | 0.020 | [-0.27 0.3 ] | 0.892 | 0.052 | 0.899 |
|  | FT_stat_Valley | 0.134 | [-0.16 0.4 ] | 0.364 | 0.149 | 0.523 |
|  | loudness_sma3_stddevRisingSlope | 0.177 | [-0.11 0.44] | 0.229 | 0.227 | 0.575 |
|  | TrajPhonZ | -0.110 | [-0.38 0.18] | 0.456 | 0.116 | 0.959 |
